## Supplementary material for "Can alcohol consumption in Germany be reduced by alcohol screening, brief intervention and referral to treatment in primary health care? Results of a simulation study": S1 Table

S1 Table. Alcohol consumption admission probability in the German adult population in 2009, by sex, age group and educational level

| **Sex** | **Age** | **Education** | **LA** | **CD** | **HED among drinkers** | **Mean drinking levels** | **% medium risk** | **% high risk** |
| --- | --- | --- | --- | --- | --- | --- | --- | --- |
| female | 15-34 | low | 11.8%  (5.9 to 21.2%) | 81.4%  (73.0 to 89.2%) | 43.4%  (30.5 to 55.9%) | 21.6  (20.7 to 23.1) | 18.1%  (10.3 to 27.4%) | 16.3%  (11.9 to 20.9%) |
|  |  | medium | 2.9%  (0.5 to 8.8%) | 89.1%  (83.1 to 94.6%) | 33.4%  (24.9 to 39.4%) | 15.3  (14.9 to 15.7) | 16.1%  ( 8.9 to 22.6%) | 9.1%  ( 5.5 to 12.2%) |
|  |  | high | 3.1%  (0.0 to 7.1%) | 90.1%  (83.2 to 95.1%) | 26.7%  (18.1 to 35.0%) | 10.5  (10.1 to 10.9) | 11.8%  ( 6.1 to 16.0%) | 3.5%  ( 1.1 to 6.8%) |
|  | 35-49 | low | 10.6%  (3.5 to 20.1%) | 71.1%  (58.1 to 82.1%) | 9.7%  ( 1.9 to 17.0%) | 3.1  ( 2.8 to 3.5) | 0.0%  ( 0.0 to 2.2%) | 0.0%  ( 0.0 to 0.0%) |
|  |  | medium | 3.8%  (1.0 to 7.7%) | 86.8%  (79.0 to 93.9%) | 12.9%  ( 6.0 to 22.0%) | 5.0  ( 4.8 to 5.1) | 2.4%  ( 0.0 to 4.8%) | 0.0%  ( 0.0 to 1.2%) |
|  |  | high | 1.0%  (0.0 to 4.1%) | 92.1%  (87.1 to 97.0%) | 13.4%  ( 6.3 to 20.9%) | 5.8  ( 5.6 to 6.0) | 4.4%  ( 1.6 to 7.7%) | 0.0%  ( 0.0 to 2.2%) |
|  | 50-64 | low | 11.4%  (4.7 to 19.7%) | 71.1%  (60.2 to 80.7%) | 9.1%  ( 2.5 to 20.2%) | 3.1  ( 2.8 to 3.4) | 0.0%  ( 0.0 to 3.0%) | 0.0%  ( 0.0 to 0.0%) |
|  |  | medium | 1.9%  (0.0 to 4.7%) | 84.7%  (74.5 to 91.9%) | 10.2%  ( 3.8 to 18.2%) | 4.4  ( 4.2 to 4.6) | 1.2%  ( 0.0 to 3.6%) | 0.0%  ( 0.0 to 0.6%) |
|  |  | high | 0.0%  (0.0 to 1.9%) | 92.1%  (86.1 to 97.0%) | 9.7%  ( 4.3 to 16.0%) | 4.6  ( 4.4 to 4.7) | 2.2%  ( 0.0 to 4.6%) | 0.0%  ( 0.0 to 1.1%) |
|  | 65-99 | low | 11.8%  (4.8 to 17.7%) | 76.7%  (65.1 to 84.8%) | 25.8%  (17.9 to 36.5%) | 10.8  (10.2 to 11.4) | 12.2%  ( 5.1 to 17.4%) | 3.8%  ( 1.3 to 6.5%) |
|  |  | medium | 2.9%  (0.0 to 6.8%) | 86.7%  (79.2 to 93.1%) | 19.0%  (10.8 to 27.2%) | 7.7  ( 7.6 to 7.9) | 7.6%  ( 3.5 to 12.2%) | 1.2%  ( 0.0 to 3.6%) |
|  |  | high | 1.0%  (0.0 to 4.2%) | 91.9%  (84.3 to 96.5%) | 16.3%  ( 7.8 to 21.2%) | 6.8  ( 6.7 to 6.9) | 5.8%  ( 2.7 to 10.6%) | 1.1%  ( 0.0 to 2.3%) |
| male | 15-34 | low | 10.0%  (4.0 to 20.0%) | 85.1%  (73.8 to 93.1%) | 66.7%  (58.6 to 75.9%) | 65.7  (63.2 to 70.2) | 12.6%  ( 6.6 to 21.3%) | 37.4%  (29.7 to 44.8%) |
|  |  | medium | 2.2%  (0.0 to 6.5%) | 91.9%  (86.1 to 97.0%) | 61.5%  (52.6 to 68.3%) | 56.7  (55.4 to 58.2) | 12.2%  ( 6.6 to 18.8%) | 33.3%  (27.0 to 38.9%) |
|  |  | high | 1.1%  (0.0 to 5.6%) | 94.6%  (89.3 to 98.1%) | 57.8%  (49.2 to 66.4%) | 45.8  (44.5 to 47.4) | 11.5%  ( 6.2 to 19.1%) | 26.5%  (21.2 to 31.3%) |
|  | 35-49 | low | 3.4%  (0.0 to 9.5%) | 73.3%  (61.3 to 85.2%) | 55.3%  (46.3 to 67.3%) | 66.6  (59.2 to 73.8) | 11.1%  ( 5.8 to 18.2%) | 37.1%  (27.9 to 45.2%) |
|  |  | medium | 2.0%  (0.0 to 6.0%) | 92.2%  (85.4 to 97.1%) | 46.1%  (37.2 to 55.9%) | 42.9  (41.1 to 44.7) | 11.6%  ( 6.4 to 18.6%) | 23.4%  (18.8 to 29.2%) |
|  |  | high | 1.0%  (0.0 to 3.0%) | 94.2%  (89.3 to 98.1%) | 36.0%  (27.0 to 45.1%) | 25.2  (24.7 to 25.8) | 9.4%  ( 4.3 to 14.5%) | 10.7%  ( 7.6 to 15.2%) |
|  | 50-64 | low | 3.7%  (0.0 to 10.7%) | 73.1%  (59.8 to 85.9%) | 56.3%  (46.1 to 68.0%) | 66.4  (58.1 to 74.8) | 11.6%  ( 4.8 to 20.0%) | 37.5%  (30.1 to 48.1%) |
|  |  | medium | 1.2%  (0.0 to 4.6%) | 87.7%  (77.5 to 94.5%) | 43.9%  (33.0 to 53.7%) | 41.9  (40.2 to 44.0) | 11.3%  ( 5.5 to 18.4%) | 23.6%  (18.0 to 29.4%) |
|  |  | high | 0.0%  (0.0 to 2.1%) | 92.9%  (85.7 to 98.0%) | 41.4%  (31.1 to 49.8%) | 40.8  (39.6 to 42.1) | 12.6%  ( 5.5 to 19.4%) | 23.0%  (17.6 to 27.0%) |
|  | 65-99 | low | 7.8%  (2.9 to 12.8%) | 80.0%  (71.7 to 88.3%) | 62.9%  (54.3 to 71.3%) | 67.1  (63.0 to 70.5) | 11.4%  ( 5.2 to 19.5%) | 37.7%  (30.6 to 45.8%) |
|  |  | medium | 2.0%  (0.0 to 5.0%) | 90.6%  (84.9 to 95.9%) | 50.2%  (41.6 to 59.8%) | 47.6  (46.5 to 48.7) | 10.9%  ( 6.0 to 17.3%) | 26.8%  (22.6 to 33.9%) |
|  |  | high | 1.0%  (0.0 to 4.0%) | 94.0%  (89.2 to 97.6%) | 42.2%  (33.1 to 49.9%) | 34.5  (33.9 to 35.1) | 12.0%  ( 5.8 to 16.4%) | 18.1%  (13.7 to 22.8%) |
