## Supplementary material for "Can alcohol consumption in Germany be reduced by alcohol screening, brief intervention and referral to treatment in primary health care? Results of a simulation study": S1 File: SBIRT impact_1_prepare data.R

```
# ========================================================================================================================================================================================================
# ========================================================================================================================================================================================================
# ========================================================================================================================================================================================================

#	CODE FOR MANUSCRIPT: 
# Can alcohol consumption in Germany be reduced by alcohol screening, brief intervention and referral to treatment in primary health care? Results of a simulation study

# Author of code:
# Jakob Manthey

# ========================================================================================================================================================================================================
# ========================================================================================================================================================================================================
# ========================================================================================================================================================================================================
# ========================================================================================================================================================================================================
# ========================================================================================================================================================================================================
# ========================================================================================================================================================================================================

# _____________________________________________________________________
#	LOAD ALL NECESSARY LIBRARIES:
# _____________________________________________________________________

# ................................................................................
#	LIBRARIES
# ................................................................................

library (data.table)
library (Hmisc)
library (openxlsx)
library (MASS)
library (stringr)
library (dglm)

# ========================================================================================================================================================================================================
# ========================================================================================================================================================================================================
# ========================================================================================================================================================================================================
# ========================================================================================================================================================================================================
# ========================================================================================================================================================================================================
# ========================================================================================================================================================================================================

# _____________________________________________________________________
# PREPARE WORKSPACE:
# _____________________________________________________________________

##  1) ALL-CAUSE
# ................................................................................

setwd("C:/Data.work/Publications/PlosOne_SBIRT Germany/S1 file")

# _____________________________________________________________________
#	LOAD DATA
# _____________________________________________________________________

##  1) Load alcohol data from ESA
# ................................................................................

alc.input <- data.table(read.xlsx("INPUT.xlsx", sheet = "ESA" ))

alc.input <- alc.input[,.(year,sex,age,ses,
                           LA, LA.SE, 
                           CD, CD.SE,
                           DDL, DDL.SE,
                           HED_DRINKER, HED_DRINKER.SE,
                           HED.LOW,	HED.LOW.SE,	HED.MID,	HED.MID.SE, HED.HIGH,	HED.HIGH.SE)]

##  2) Load PHC data from GEDA
# ................................................................................

geda.input <- data.table(read.xlsx("INPUT.xlsx", sheet = "GEDA"))

phc.data <- copy(geda.input[,.(sex,age,ses,N,
                               PHC.any.alcohol, PHC.any.alcohol.SE, 
                               PHC.risk.low, PHC.risk.low.SE,
                               PHC.risk.mid, PHC.risk.mid.SE,
                               PHC.risk.high, PHC.risk.high.SE)])


##  3) Load APC data
# ................................................................................
   
apc.input <- data.table(read.xlsx("INPUT.xlsx", sheet = "APC"))
apc.data <- copy(apc.input)
apc.data$APC.SE <- 0 
apc.data[, APC.diff := c(0,diff(APC,1))]
apc.data[, APC.diff.rate := APC.diff / APC]

##  4) Load Population data
# ................................................................................

#  4.1.) UN population data (from world population prospects 2019 revision):
pop.1 <- data.table(read.csv("WPP2019_Germany.csv"))

#  4.2) EUROSTAT breakup by educational group

# downloaded on 10 Feb 2021 from Eurostat: Population by sex, age and educational attainment level (1 000) => "lfsa_pgaed.zip"

pop.2 <- data.table(read.csv("lfsa_pgaed_1_Data.csv"))
pop.2 <- pop.2[,.(year = TIME, sex = ifelse(SEX == "Males", "male", "female"),age2 = AGE,ISCED11,pop = as.numeric(gsub(",","",Value)))]
pop.2[, ses := ifelse(ISCED11 == "Less than primary, primary and lower secondary education (levels 0-2)", "low",
                       ifelse(ISCED11 == "Upper secondary and post-secondary non-tertiary education (levels 3 and 4)", "mid",
                              ifelse(ISCED11 == "Tertiary education (levels 5-8)", "high",NA)))]
pop.2[sex == "female", mean(pop), by = .(year,ses)]
pop.2[, age2 := gsub("From ", "", age2)]
pop.2[, age2 := gsub(" years", "", age2)]
pop.2[, age2 := gsub(" to ", "-", age2)]
pop.2[, unique(age2)]
pop.2[, age := ifelse(age2 %in% c("15-24","25-29","30-34"),"15-34",
                     ifelse(age2 %in% c("35-39","40-44","45-49"),"35-49",age2))]
pop.2[, unique(age)]
pop.2 <- pop.2[!is.na(age),.(pop = sum(pop)), by = .(year,sex,age,ses)]

#  get 65-74 via 50-74 minus 50-64
temp <- dcast(pop.2[age %in% c("50-74","50-64")], year + sex + ses ~ age, value.var = "pop")
names(temp) <- c("year","sex","ses","part","all")
temp <- temp[,.(year,sex,age = "65-99",ses,pop = all - part)] # label new age group as 65-99 because we assume that the educational proportions (!) are the same as in 65-74
pop.2 <- rbind(pop.2[age != "50-74"], temp)

#  get ses proportions within each year,sex,age group
pop.2[, denom := sum(pop), by = .(year,sex,age)]
pop.2[, ses.prop := pop / denom]
pop.2[, sum(ses.prop), by = .(year,sex,age)] # should be all = 1

#  combine pop.1 and pop.2
pop.data <- merge(pop.1, pop.2[,.(year,sex,age,ses,ses.prop)], by = c("year","sex","age"), all.x = T)
pop.data <- pop.data[,.(year,sex,age,ses,pop = ses.prop * pop)]

pop.data[!complete.cases(pop.data)]


# _____________________________________________________________________
#	DEFINE SOME PARAMETERS
# _____________________________________________________________________

##  1) age groups, risk thresholds
# ................................................................................

# age groups
age.groups <- c("15-34","35-49","50-64","65-99")

# thresholds
risk.threshold.1    <- c(female = 20.9999, male = 40.9999)
risk.threshold.2    <- c(female = 40.9999, male = 60.9999)

##  2) BI and RT effect sizes
# ................................................................................

##  BI - DRINKING LEVELS

##  Kaner et al 2018:
  #   weekly baseline drinking levels: females: 190; males: 350
  #   absolute reduction: females: -30 (-59 to -2); males: -42 (-65 to -20)
  #   proportional reduction: -30/190

bi.eff.fem   <- c(est = -30/190, low = -59/190, hi = -2/190)
bi.eff.fem.sd <- mean(c(bi.eff.fem[1] - bi.eff.fem[2], bi.eff.fem[3] - bi.eff.fem[1]))/1.96
bi.eff.mal   <- c(est = -42/350, low = -65/350, hi = -20/350)
bi.eff.mal.sd <- mean(c(bi.eff.mal[1] - bi.eff.mal[2], bi.eff.mal[3] - bi.eff.mal[1]))/1.96

bi.eff.dl      <- data.table(cbind(var = c("est","sd"),
                                   female = c(bi.eff.fem[1],bi.eff.fem.sd),
                                   male   = c(bi.eff.mal[1],bi.eff.mal.sd)))

##  check if effect and CI can be reproduced: 
temp <- rnorm(mean = bi.eff.fem[1], sd = bi.eff.fem.sd, n = 10000)
mean(temp) ; bi.eff.fem[1]
quantile(temp, 0.025) ; bi.eff.fem[2]
quantile(temp, 0.975) ; bi.eff.fem[3]
rm(temp, bi.eff.fem, bi.eff.fem.sd, bi.eff.mal, bi.eff.mal.sd)

##  BI - HED STATUS

##  Kaner et al 2018 ("Analysis 1.22. Comparison 1 Brief intervention versus control, Outcome 22 Binge drinkers at 12 months"):
  #  Risk difference = -0.07 [ -0.12, -0.02 ]

bi.eff.hed <- c(est = -0.07, sd = mean(c(-0.07 - -0.12, -0.02 - -0.07))/1.96)

##  RT

##  probability of intervention if screened:
rt.prob   <- 0.5
     
##  effect sizes: => same as BI
rt.eff.dl <- copy(bi.eff.dl)
rt.eff.hed <- copy(bi.eff.hed)

##  HED

 # ADD BI.effect.HED from Kaner et al 2007 ==> 11% risk reduction
 

##  3) BI distribution by SES
# ................................................................................

##  probability of intervention if screened:
interv.prob   <- 0.5

##  4) Attenuation of intervention parameters
# ................................................................................

##  main analyses: stable effects for four years (Fleming et al., 200), linear decline therafter and nullifying 10years post intervention (Wutzke et al., 2002)

attenuation.main <- data.table(year = c(1:10), attenuation = c(1,1,1,seq(1,0,length.out =  7)))
 
##  sensitivity analyses: stable effects for four years (Fleming et al., 200), linear decline therafter and nullifying 10years post intervention (Wutzke et al., 2002)

attenuation.sens <- data.table(year = c(1:10), attenuation = c(seq(1,0,length.out =  3),rep(0,7)))    

# ========================================================================================================================================================================================================
# ========================================================================================================================================================================================================
# ========================================================================================================================================================================================================# ========================================================================================================================================================================================================
# ========================================================================================================================================================================================================
# ========================================================================================================================================================================================================

# _____________________________________________________________________
# PREPARE DATA:
# _____________________________________________________________________

##  1) ALCOHOL
# ................................................................................

##  1.0): correct SEs
alc.input[LA %in% c(0,1), LA.SE := 0]
alc.input[CD %in% c(0,1), CD.SE := 0]
alc.input[HED_DRINKER %in% c(0,1), HED_DRINKER.SE := 0]
alc.input[HED.LOW %in% c(0,1), HED.LOW.SE := 0]
alc.input[HED.MID %in% c(0,1), HED.MID.SE := 0]
alc.input[HED.HIGH %in% c(0,1), HED.HIGH.SE := 0]
 
##  1.1): CREATE a consistent time series of alc data
   # combine to get time series
     year.length <- 2 * 4 * 3
     alc.data <- data.table(matrix(data = NA, nrow = (year.length * 10), ncol = 4, 
                                   dimnames = list(NULL,names(alc.input)[1:4])))
     
     alc.data$year <- rep(seq(2009,2018),each = year.length)
     alc.data$sex <- rep(rep(c("female","male"),each = year.length  / 2),10)
     alc.data$age <- rep(rep(age.groups,each = 3),20)
     alc.data$ses <- rep(c("low","mid","high"),year.length  / 3 * 10)
 
     alc.input[, age := ifelse( age == "18-34", "15-34", age)]
     alc.data <- merge(alc.data, alc.input, by = c("year","sex","age","ses"), all.x = T)
     rm(year.length)
     
## 1.2) IMPUTATION within key years
   # impute missing variables for row.
     # done manually in Excel. rules: 
     # 1 - take values of same sex and ses but neighbouring age group
     # 2 - take values of same sex and ses across all age groups
     
   # 1.2.1) impute low SES : 35-49 and 50-64 with 35-64
     var.names <- c("LA","LA.SE","CD","CD.SE","DDL","DDL.SE","HED_DRINKER","HED_DRINKER.SE","HED.LOW","HED.LOW.SE","HED.MID","HED.MID.SE","HED.HIGH","HED.HIGH.SE")
     
     # years for which alc.data is available
     key.years <- c(2009, 2012, 2015, 2018)
     
     for (y in key.years){
    
       alc.data[year == y & ses == "low" & sex == "female" & age == "35-49",c("LA","LA.SE","CD","CD.SE","DDL","DDL.SE","HED_DRINKER","HED_DRINKER.SE","HED.LOW","HED.LOW.SE","HED.MID","HED.MID.SE","HED.HIGH","HED.HIGH.SE")] <-
         alc.input[year == y & ses == "low" & sex == "female" & age == "35-64",c("LA","LA.SE","CD","CD.SE","DDL","DDL.SE","HED_DRINKER","HED_DRINKER.SE","HED.LOW","HED.LOW.SE","HED.MID","HED.MID.SE","HED.HIGH","HED.HIGH.SE")]
        
      alc.data[year == y & ses == "low" & sex == "female" & age == "50-64",c("LA","LA.SE","CD","CD.SE","DDL","DDL.SE","HED_DRINKER","HED_DRINKER.SE","HED.LOW","HED.LOW.SE","HED.MID","HED.MID.SE","HED.HIGH","HED.HIGH.SE")] <- 
        alc.input[year == y & ses == "low" & sex == "female" & age == "35-64",c("LA","LA.SE","CD","CD.SE","DDL","DDL.SE","HED_DRINKER","HED_DRINKER.SE","HED.LOW","HED.LOW.SE","HED.MID","HED.MID.SE","HED.HIGH","HED.HIGH.SE")]
            
      alc.data[year == y & ses == "low" & sex == "male" & age == "35-49",c("LA","LA.SE","CD","CD.SE","DDL","DDL.SE","HED_DRINKER","HED_DRINKER.SE","HED.LOW","HED.LOW.SE","HED.MID","HED.MID.SE","HED.HIGH","HED.HIGH.SE")] <- 
        alc.input[year == y & ses == "low" & sex == "male" & age == "35-64",c("LA","LA.SE","CD","CD.SE","DDL","DDL.SE","HED_DRINKER","HED_DRINKER.SE","HED.LOW","HED.LOW.SE","HED.MID","HED.MID.SE","HED.HIGH","HED.HIGH.SE")]
      
      alc.data[year == y & ses == "low" & sex == "male" & age == "50-64",c("LA","LA.SE","CD","CD.SE","DDL","DDL.SE","HED_DRINKER","HED_DRINKER.SE","HED.LOW","HED.LOW.SE","HED.MID","HED.MID.SE","HED.HIGH","HED.HIGH.SE")] <-
        alc.input[year == y & ses == "low" & sex == "male" & age == "35-64",c("LA","LA.SE","CD","CD.SE","DDL","DDL.SE","HED_DRINKER","HED_DRINKER.SE","HED.LOW","HED.LOW.SE","HED.MID","HED.MID.SE","HED.HIGH","HED.HIGH.SE")]
        
     }
     
      alc.data[year %in% key.years & is.na(LA) & age != "65-99"] # should be none
      alc.data[year %in% key.years & is.na(LA.SE) & age != "65-99"] # should be none
      alc.data[year %in% key.years & is.na(CD) & age != "65-99"] # should be none
      alc.data[year %in% key.years & is.na(CD.SE) & age != "65-99"] # should be none
      alc.data[year %in% key.years & is.na(HED_DRINKER) & age != "65-99"] # should be none
      alc.data[year %in% key.years & is.na(HED_DRINKER.SE) & age != "65-99"] # should be none

    # 1.2.2) use sex-ses specific HED RISK - not age specific to avoid low N / high SE
      for (y in key.years){
        
        alc.data[year == y & ses == "low" & sex == "female" & age != "65-99",c("HED.LOW","HED.LOW.SE","HED.MID","HED.MID.SE","HED.HIGH","HED.HIGH.SE")] <-
          alc.input[year == y & ses == "low" & sex == "female" & age %like% "18-64",c("HED.LOW","HED.LOW.SE","HED.MID","HED.MID.SE","HED.HIGH","HED.HIGH.SE")]
          
        alc.data[year == y & ses == "low" & sex == "male" & age != "65-99",c("HED.LOW","HED.LOW.SE","HED.MID","HED.MID.SE","HED.HIGH","HED.HIGH.SE")] <-
          alc.input[year == y & ses == "low" & sex == "male" & age %like% "18-64",c("HED.LOW","HED.LOW.SE","HED.MID","HED.MID.SE","HED.HIGH","HED.HIGH.SE")]
      }
      
      alc.data[year %in% key.years & is.na(HED.LOW) & age != "65-99"] # should be none
      alc.data[year %in% key.years & is.na(HED.LOW.SE) & age != "65-99"] # should be none
      alc.data[year %in% key.years & is.na(HED.MID) & age != "65-99"] # should be none
      alc.data[year %in% key.years & is.na(HED.MID.SE) & age != "65-99"] # should be none
      alc.data[year %in% key.years & is.na(HED.HIGH) & age != "65-99"] # should be none
      alc.data[year %in% key.years & is.na(HED.HIGH.SE) & age != "65-99"] # should be none
      
      
    # 1.2.3) check if there's any SE > .2
      
      alc.data[, .SD, .SDcols = names(alc.data) %like% "LA.SE|CD.SE"]
      alc.data[LA.SE > .2] # none
      alc.data[CD.SE > .2] # none
      alc.data[, summary(LA.SE)] # max .04
      alc.data[, summary(CD.SE)] # max .05
      
    # 1.2.4) use total pop for estimate for 65+
      
      for (y in key.years){
        
        alc.data[year == y & sex == "female" & age == "65-99" & ses == "low",c("LA","LA.SE","CD","CD.SE","DDL","DDL.SE","HED_DRINKER","HED_DRINKER.SE","HED.LOW","HED.LOW.SE","HED.MID","HED.MID.SE","HED.HIGH","HED.HIGH.SE")] <- 
          alc.input[year == y & sex == "female" & age == "18-64" & ses == "low",c("LA","LA.SE","CD","CD.SE","DDL","DDL.SE","HED_DRINKER","HED_DRINKER.SE","HED.LOW","HED.LOW.SE","HED.MID","HED.MID.SE","HED.HIGH","HED.HIGH.SE")]
        
        alc.data[year == y & sex == "female" & age == "65-99" & ses == "mid",c("LA","LA.SE","CD","CD.SE","DDL","DDL.SE","HED_DRINKER","HED_DRINKER.SE","HED.LOW","HED.LOW.SE","HED.MID","HED.MID.SE","HED.HIGH","HED.HIGH.SE")] <- 
          alc.input[year == y & sex == "female" & age == "18-64" & ses == "mid",c("LA","LA.SE","CD","CD.SE","DDL","DDL.SE","HED_DRINKER","HED_DRINKER.SE","HED.LOW","HED.LOW.SE","HED.MID","HED.MID.SE","HED.HIGH","HED.HIGH.SE")]
        
        alc.data[year == y & sex == "female" & age == "65-99" & ses == "high",c("LA","LA.SE","CD","CD.SE","DDL","DDL.SE","HED_DRINKER","HED_DRINKER.SE","HED.LOW","HED.LOW.SE","HED.MID","HED.MID.SE","HED.HIGH","HED.HIGH.SE")] <- 
          alc.input[year == y & sex == "female" & age == "18-64" & ses == "high",c("LA","LA.SE","CD","CD.SE","DDL","DDL.SE","HED_DRINKER","HED_DRINKER.SE","HED.LOW","HED.LOW.SE","HED.MID","HED.MID.SE","HED.HIGH","HED.HIGH.SE")]
      
        alc.data[year == y & sex == "male" & age == "65-99" & ses == "low",c("LA","LA.SE","CD","CD.SE","DDL","DDL.SE","HED_DRINKER","HED_DRINKER.SE","HED.LOW","HED.LOW.SE","HED.MID","HED.MID.SE","HED.HIGH","HED.HIGH.SE")] <- 
          alc.input[year == y & sex == "male" & age == "18-64" & ses == "low",c("LA","LA.SE","CD","CD.SE","DDL","DDL.SE","HED_DRINKER","HED_DRINKER.SE","HED.LOW","HED.LOW.SE","HED.MID","HED.MID.SE","HED.HIGH","HED.HIGH.SE")]
        
        alc.data[year == y & sex == "male" & age == "65-99" & ses == "mid",c("LA","LA.SE","CD","CD.SE","DDL","DDL.SE","HED_DRINKER","HED_DRINKER.SE","HED.LOW","HED.LOW.SE","HED.MID","HED.MID.SE","HED.HIGH","HED.HIGH.SE")] <- 
          alc.input[year == y & sex == "male" & age == "18-64" & ses == "mid",c("LA","LA.SE","CD","CD.SE","DDL","DDL.SE","HED_DRINKER","HED_DRINKER.SE","HED.LOW","HED.LOW.SE","HED.MID","HED.MID.SE","HED.HIGH","HED.HIGH.SE")]
        
        alc.data[year == y & sex == "male" & age == "65-99" & ses == "high",c("LA","LA.SE","CD","CD.SE","DDL","DDL.SE","HED_DRINKER","HED_DRINKER.SE","HED.LOW","HED.LOW.SE","HED.MID","HED.MID.SE","HED.HIGH","HED.HIGH.SE")] <- 
          alc.input[year == y & sex == "male" & age == "18-64" & ses == "high",c("LA","LA.SE","CD","CD.SE","DDL","DDL.SE","HED_DRINKER","HED_DRINKER.SE","HED.LOW","HED.LOW.SE","HED.MID","HED.MID.SE","HED.HIGH","HED.HIGH.SE")]

      }


# 1.3) IMPUTATION across missing years:
    
    # function
    impute <- function(v1, v2, len) {
      diff <- v2 - v1
      step <- diff / (len -1)
      out <- v1
      if(v1 == 0 & is.na(v2)){
        out <- rep(0, len) 
      } else {
        i <- 1
        while (i < len) {
          out <- c(out, out[length(out)] + step)
          i <- i + 1
        }  
      }
      
      return(out)
    }

    # test:
    alc.data[year %in% c(2009:2012) & sex == "female" & age == "15-34" & ses == "low", .(year,HED.HIGH)]
    impute(v1 = 1, v2 = 0.969755, len = 4)
    impute(v1 = 1, v2 = 1, len = 4)
    impute(v1 = 0, v2 = 0, len = 4)
    
    # check and apply
    alc.data[year %in% key.years][!complete.cases(alc.data[year %in% key.years])] # should be no missing values in key years
    
    y <- 1
    while(y < length(key.years)){
      
      alc.data[year %in% c(key.years[y]:key.years[y+1]), ':=' (
        LA     = impute(v1 = LA[1]    , v2 = LA[.N]    , len = .N),
        LA.SE  = impute(v1 = LA.SE[1] , v2 = LA.SE[.N] , len = .N),
        CD     = impute(v1 = CD[1]    , v2 = CD[.N]    , len = .N),
        CD.SE  = impute(v1 = CD.SE[1] , v2 = CD.SE[.N] , len = .N),
        DDL    = impute(v1 = DDL[1]   , v2 = DDL[.N]   , len = .N),
        DDL.SE = impute(v1 = DDL.SE[1], v2 = DDL.SE[.N], len = .N),
        HED_DRINKER     = impute(v1 = HED_DRINKER[1]   , v2 = HED_DRINKER[.N]   , len = .N),
        HED_DRINKER.SE  = impute(v1 = HED_DRINKER.SE[1], v2 = HED_DRINKER.SE[.N], len = .N),
        HED.LOW     = impute(v1 = HED.LOW[1]   , v2 = HED.LOW[.N]   , len = .N),
        HED.LOW.SE  = impute(v1 = HED.LOW.SE[1], v2 = HED.LOW.SE[.N], len = .N),
        HED.MID     = impute(v1 = HED.MID[1]   , v2 = HED.MID[.N]   , len = .N),
        HED.MID.SE  = impute(v1 = HED.MID.SE[1], v2 = HED.MID.SE[.N], len = .N),
        HED.HIGH    = impute(v1 = HED.HIGH[1]   , v2 = HED.HIGH[.N]   , len = .N),
        HED.HIGH.SE = impute(v1 = HED.HIGH.SE[1], v2 = HED.HIGH.SE[.N], len = .N)
        ), 
      by = .(sex,age,ses)]
      
      y <- y+1
      
    }
    
    # check
    alc.data[year <= 2018 & sex == "female" & age == "15-34" & ses == "low",.(year,sex,age,ses,LA,LA.SE)]
    alc.data[year <= 2018 & sex == "female" & age == "15-34" & ses == "low",.(year,sex,age,ses,CD,CD.SE)]
    alc.data[year <= 2018 & sex == "female" & age == "15-34" & ses == "low",.(year,sex,age,ses,DDL,DDL.SE)]
    alc.data[year <= 2018 & sex == "female" & age == "15-34" & ses == "low",.(year,sex,age,ses,HED_DRINKER,HED_DRINKER.SE)]
    alc.data[year <= 2018 & sex == "female" & age == "15-34" & ses == "low",.(year,sex,age,ses,HED.LOW,HED.LOW.SE)]
    alc.data[year <= 2018 & sex == "female" & age == "15-34" & ses == "low",.(year,sex,age,ses,HED.MID,HED.MID.SE)]
    alc.data[year <= 2018 & sex == "female" & age == "15-34" & ses == "low",.(year,sex,age,ses,HED.HIGH,HED.HIGH.SE)]
    alc.data[year <= 2018 & sex == "female" & age == "15-34" & ses == "high",.(year,sex,age,ses,HED.HIGH,HED.HIGH.SE)]
    alc.data[year %in% c(key.years) ,.(year,sex,age,ses,HED.HIGH)]
    
    # check LA+CD
    alc.data[LA + CD > 1,.(year,sex,age,ses,LA,CD)] #none

    # check any missing values?
    alc.data[!complete.cases(alc.data)] # none

# 1.4) ADD APC
    
    alc.data <- merge(alc.data, apc.data[,.(year, APC.total = APC, APC.SE)], by = "year", all.x = T)
    alc.data[!complete.cases(alc.data)] # should be none
    
    # add population
    alc.data <- merge(alc.data, pop.data, by = c("year","sex","age","ses"), all.x = T)
    alc.data[!complete.cases(alc.data)] # should be none
    
    # plot
    ggplot(alc.data, aes(age, DDL, color = ses)) + facet_grid(sex ~ .) + geom_point()
    ggplot(alc.data, aes(age, HED_DRINKER, color = ses)) + facet_grid(sex ~ .) + geom_point()
    
    # split APC by sex, age, ses
    
        # first: determine drinking volume per sex-age-ses group and relative proportion
        #alc.data[, dl := drinking.level * CD * pop]
        alc.data[, dl := DDL * CD * HED_DRINKER * pop]
        alc.data[, dl.total := sum(dl), by = year]
        alc.data[, dl_prop := dl / dl.total]
        
        # second: rescale APC with these proportions:
        alc.data[, DL.total := APC.total * sum(pop), by = year]
        alc.data[, DL := DL.total * dl_prop]
        alc.data[, APC := DL / pop]
        
        # third: check
        round(alc.data[year == 2009, wtd.mean(APC, pop)],3) == round(apc.data[year == 2009]$APC,3)
        round(alc.data[year == 2012, wtd.mean(APC, pop)],3) == round(apc.data[year == 2012]$APC,3)
        round(alc.data[year == 2015, wtd.mean(APC, pop)],3) == round(apc.data[year == 2015]$APC,3)
        round(alc.data[year == 2018, wtd.mean(APC, pop)],3) == round(apc.data[year == 2018]$APC,3)
    
    # fourth: clean
    alc.data <- alc.data[, .SD, .SDcols = c(names(alc.input),"APC","APC.SE","pop")]
    alc.data[!complete.cases(alc.data)] # should be none
    
# 1.5) RESCALE HED probabilities:
    
    # a) get % of drinkers in each category
    alc.data2 <- copy(alc.data[,.(year,sex,age,ses,
                                  APC.DRINKER = APC / CD,
                                  P_real = HED_DRINKER,
                                  p1 = HED.LOW,
                                  p2 = HED.MID,
                                  p3 = HED.HIGH)])
    
    alc.data2[, ':=' ( GAMMA.MEAN = APC.DRINKER * 1000 * 0.789 / 365)]
    alc.data2[, ':=' ( GAMMA.SD   = ifelse(sex == "male", 1.171 * GAMMA.MEAN , 1.258 * GAMMA.MEAN))]
    alc.data2[, ':=' ( KAPPA = GAMMA.MEAN^2 / GAMMA.SD^2,
                       THETA = GAMMA.SD^2 / GAMMA.MEAN)]
    alc.data2[, ':=' ( w1 = integrate(dgamma, lower = 0.00001,               upper = risk.threshold.1[sex], shape = KAPPA, scale = THETA )$value,
                       w2 = integrate(dgamma, lower = risk.threshold.1[sex], upper = risk.threshold.2[sex], shape = KAPPA, scale = THETA )$value,
                       w3 = integrate(dgamma, lower = risk.threshold.2[sex], upper = 150                  , shape = KAPPA, scale = THETA )$value), 
              by = .(year,sex,age,ses)]
    alc.data2[, ':=' ( total = rowSums(alc.data2[,.(w1,w2,w3)]) )]
    alc.data2[, ':=' ( w1 = w1 / total ,
                       w2 = w2 / total ,
                       w3 = w3 / total ) ]
    alc.data2$total <- NULL
  
    # b) rescale HED probability for medium risk drinkers:
      # formula: 
      # P_real = P_alt * x1 = (p1*w1*x2) + (p2*w2) + (p3*w3)
      # x2=(P_real-p2*w2-p3*w3)/(p1*w1)
      
      alc.data2[, ':=' ( P_alt = sum(c(p1 * w1,
                                       p2 * w2,
                                       p3 * w3) )), by = .(year,sex,age,ses)]
      
      alc.data2[, ':=' ( x1 = P_real / P_alt )]
      
      alc.data2[, ':=' ( P_recalc = sum(c(p1 * w1,
                                          p2 * w2,
                                          p3 * w3) * x1 )), by = .(year,sex,age,ses)]
      
      alc.data2[,.(year,sex,age,ses,P_real,x1,P_alt,P_recalc)] # ok
      
      alc.data2[, ':=' ( x2 = ((P_real)-(p2 * w2)-(p3 * w3))/(p1 * w1)),
                by = .(year,sex,age,ses)]
      
      alc.data2[, ':=' ( P_recalc = sum(c(p1 * w1 * x2,
                                          p2 * w2,
                                          p3 * w3))), by = .(year,sex,age,ses)]
      
      alc.data2[,.(year,sex,age,ses,P_real,P_recalc)] # ok
      alc.data2[,all(round(P_real,3) == round(P_recalc,3))] # should be T
    
      # adjust probability
      alc.data <- merge(alc.data, alc.data2[,.(year,sex,age,ses,HED.LOW.ADJUST = p1 * x2)], by = c("year","sex","age","ses"), all.x = T)
      ggplot( melt(alc.data[, .(year,sex,age,ses,HED.LOW,HED.LOW.ADJUST)], id.vars = c("year","sex","age","ses")), aes(x = year, y = value, color = variable) ) + geom_point()
      ggplot( alc.data[, .(year,sex,age,ses,diff=HED.LOW-HED.LOW.ADJUST)], aes(x = year, y = diff) ) + geom_point() 
      # in most observations lower than reported, on average = 16%
      alc.data[, .(mean(HED.LOW-HED.LOW.ADJUST))]
      alc.data[, summary(HED.LOW.ADJUST)]
      alc.data[, HED.LOW := HED.LOW.ADJUST]
      alc.data$HED.LOW.ADJUST <- NULL
      rm(alc.data2)

##  2) PHC
# ................................................................................

# correct PHC visit for high risk drinker, if cells < 100 OR NA OR = 0
phc.data[N<100 | is.na(PHC.risk.high) | PHC.risk.high == 0, ':=' (PHC.risk.high = PHC.any.alcohol,
                                                                  PHC.risk.high.SE = PHC.any.alcohol.SE)]
phc.data

phc.data <- phc.data[,.(sex,age,ses,
                        PHC.risk.low, PHC.risk.low.SE,
                        PHC.risk.mid, PHC.risk.mid.SE,
                        PHC.risk.high, PHC.risk.high.SE)]

##  3) BASE = ALC & PHC
# ................................................................................

base.data   <- merge(alc.data, phc.data, by = c("sex","age","ses"), all.x = T)

##  Clean input variables:
base.data[, age := factor(age, levels = age.groups, ordered = T)]
base.data[, ses := factor(ses, levels = c("low","mid","high"), ordered = T)]


##  4) Datatable specifying different scenarios
# ................................................................................

main.analyses <- rbind(data.table(year = c(1:10), scenario = 0, coverage = 0.029),
                       data.table(year = c(1:10), scenario = 1, coverage = 0.00),
                       data.table(year = c(1:10), scenario = 2, coverage = 0.1),
                       data.table(year = c(1:10), scenario = 3, coverage = 0.25),
                       data.table(year = c(1:10), scenario = 4, coverage = 0.5),
                       data.table(year = c(1:10), scenario = 5, coverage = 0.75))

# ========================================================================================================================================================================================================
# ========================================================================================================================================================================================================
# ========================================================================================================================================================================================================
# ========================================================================================================================================================================================================
# ========================================================================================================================================================================================================
# ========================================================================================================================================================================================================
```
