## Supplementary material for "Can alcohol consumption in Germany be reduced by alcohol screening, brief intervention and referral to treatment in primary health care? Results of a simulation study": S1 File: SBIRT impact_2_simulation.R

```
# ========================================================================================================================================================================================================
# ========================================================================================================================================================================================================
# ========================================================================================================================================================================================================

#	CODE FOR MANUSCRIPT: 
# Can alcohol consumption in Germany be reduced by alcohol screening, brief intervention and referral to treatment in primary health care? Results of a simulation study

# Author of code:
# Jakob Manthey

# ========================================================================================================================================================================================================
# ========================================================================================================================================================================================================
# ========================================================================================================================================================================================================
# ========================================================================================================================================================================================================
# ========================================================================================================================================================================================================
# ========================================================================================================================================================================================================

# _____________________________________________________________________
#	LOAD ALL NECESSARY LIBRARIES:
# _____________________________________________________________________

# ................................................................................
#	LIBRARIES
# ................................................................................

library (data.table)
library (Hmisc)
library (openxlsx)
library (MASS)
library (stringr)
library (dglm)

# ========================================================================================================================================================================================================
# ========================================================================================================================================================================================================
# ========================================================================================================================================================================================================
# ========================================================================================================================================================================================================
# ========================================================================================================================================================================================================
# ========================================================================================================================================================================================================

# _____________________________________________________________________
#	SIMULATION
# _____________________________________________________________________

##  1) DEFINE PARAMETERS
# ................................................................................

##  Function parameters:

real <- T # set to TRUE when debugging is done

if(real != T){
  
  aProb.intervention  = interv.prob
  
  aEff.bi.dl          = bi.eff.dl
  aEff.rt.dl          = rt.eff.dl
  aEff.bi.hed         = bi.eff.hed
  aEff.rt.hed         = rt.eff.hed
  
  aAttenuate = attenuation.main
  aAttenuate = attenuation.sens
  
  N.sample = 100; 
  
  aSeed = 46483
  i <- s <- 1
  CI.sample = 100
  
  data.in = copy(base.data)
  aScenario = main.analyses[scenario == 0]
  years = 10
  years.lab = c(2009:2018)
  
}


##  2) Function
#   ::::::::::::::::::::::::::::::::::::::::: 

FUN_SIM_ <- function(data.in = copy(base.data), N.sample = 100, CI.sample = 100, 
                     aScenario = main.analyses, years = 10, years.lab = c(2009:2018), 
                     aSeed = 99854121, aAttenuate = attenuation.main, 
                     aProb.intervention = interv.prob,
                     aEff.bi.dl          = bi.eff.dl,
                     aEff.rt.dl          = rt.eff.dl,
                     aEff.bi.hed         = bi.eff.hed,
                     aEff.rt.hed         = rt.eff.hed
                     ){
  
  #  A) get vectors for simulating data:
  #  ...................................
  
  vLENGTH   <- nrow(data.in[year == years.lab[1]]) * N.sample * CI.sample
  
  vSEX     <- character(length = vLENGTH*1.1)
  vAGE     <- character(length = vLENGTH*1.1)
  vSES     <- character(length = vLENGTH*1.1)
  
  vAPC.BASE<- double(length = vLENGTH*1.1)
  vAPC.BASE.DRINKER <- double(length = vLENGTH*1.1)
  
  vLA.ref <- logical(length = vLENGTH*1.1)
  vCD.ref <- logical(length = vLENGTH*1.1)
  vFD.ref <- logical(length = vLENGTH*1.1)
  
  vDL.ref  <- double(length = vLENGTH*1.1)
  vHED.ref <- logical(length = vLENGTH*1.1)
  vSIM     <- double(length = vLENGTH*1.1)

  
  #  C) Simulate Baseline data:
  #  ...................................
  
  print("... Simulating baseline alcohol consumption data ...")
  
  # Calculate all year-1 parameters

  #row <- 19
  #aSIM  <- 1
  set.seed(aSeed)
  select <- 0
  for(row in 1:nrow(data.in[year == years.lab[1]])){ #
    
    ##  determine Kappa and Theta from baseline APC
    gamma.dat <- copy( data.in[year == years.lab[1]][row,.(sex,age,ses,APC,APC.SE,
                                                           LA,LA.SE,
                                                           CD,CD.SE,
                                                           HED.LOW,HED.LOW.SE,
                                                           HED.MID,HED.MID.SE,
                                                           HED.HIGH,HED.HIGH.SE)] )
    
    # draw APC, LA and CD (these are the input values used to draw drinking levels and drinking status for each drinker!)
      APC.draw  <- gamma.dat[,  rnorm(n = CI.sample, mean = APC, sd = APC.SE)]
      APC.draw  <- APC.draw * (gamma.dat$APC/mean(APC.draw))
      
      LA.draw   <- gamma.dat[,  rnorm(n = CI.sample, mean = LA, sd = LA.SE)]
      LA.draw   <- ifelse(LA.draw < 0, 0, LA.draw)
      LA.draw   <- LA.draw * (gamma.dat$LA/mean(LA.draw))
      
      CD.draw   <- gamma.dat[,  rnorm(n = CI.sample, mean = CD, sd = CD.SE)]
      i <- 1
      while(gamma.dat$CD != mean(CD.draw) & i < 100){
        CD.draw   <- CD.draw * (gamma.dat$CD/mean(CD.draw))
        CD.draw   <- ifelse(CD.draw > 1, 0.999, CD.draw)  
        i <- i + 1
      }
      
    # make sure that CD + LA do not exceed 1:
      adjust <- ifelse(CD.draw + LA.draw > 1, 1 / (CD.draw + LA.draw) * 0.999, 1)
      CD.draw <- CD.draw * adjust
      LA.draw <- LA.draw * adjust
      rm(adjust)
    
    # calculate APC.DRINKER  
      APC.DRINKER.draw <- APC.draw / CD.draw
      
    # distribution of HED risk by drinking level, corrected for [0,1] range
      HED.draw.risk.low   <- round(gamma.dat[,  rnorm(n = CI.sample, mean = HED.LOW, sd = HED.LOW.SE)],5)
      i <- 1
      while(round(gamma.dat$HED.LOW,4) != round(mean(HED.draw.risk.low),4) & i < 100){
        HED.draw.risk.low   <- HED.draw.risk.low * (gamma.dat$HED.LOW/mean(HED.draw.risk.low))
        HED.draw.risk.low   <- ifelse(HED.draw.risk.low < 0, 0, ifelse(HED.draw.risk.low > 1, 0.999, HED.draw.risk.low))
        i <- i + 1
      }
      
      HED.draw.risk.mid   <- round(gamma.dat[,  rnorm(n = CI.sample, mean = HED.MID, sd = HED.MID.SE)],5)
      i <- 1
      while(round(gamma.dat$HED.MID,4) != round(mean(HED.draw.risk.mid),4) & i < 100){
        HED.draw.risk.mid   <- HED.draw.risk.mid * (gamma.dat$HED.MID/mean(HED.draw.risk.mid))
        HED.draw.risk.mid   <- ifelse(HED.draw.risk.mid < 0, 0, ifelse(HED.draw.risk.mid > 1, 0.999, HED.draw.risk.mid))
        i <- i + 1
      }
      
      HED.draw.risk.high  <- round(gamma.dat[,  rnorm(n = CI.sample, mean = HED.HIGH, sd = HED.HIGH.SE)],5)
      i <- 1
      while(round(gamma.dat$HED.HIGH,4) != round(mean(HED.draw.risk.high),4) & i < 100){
        HED.draw.risk.high  <- HED.draw.risk.high * (gamma.dat$HED.HIGH/mean(HED.draw.risk.high))
        HED.draw.risk.high  <- ifelse(HED.draw.risk.high < 0, 0, ifelse(HED.draw.risk.high > 1, 0.999, HED.draw.risk.high))
        i <- i + 1
      }
      
    # calculate mean & SD from APC
      GAMMA.MEAN  <- APC.DRINKER.draw * 1000 * 0.789 / 365  ## mean g per day
      GAMMA.SD    <- ifelse(gamma.dat$sex == "male", 1.171 * GAMMA.MEAN , 1.258 * GAMMA.MEAN)
    
    # Get Kappa (shape) & Theta (scale):
      KAPPA <- GAMMA.MEAN^2 / GAMMA.SD^2
      THETA <- GAMMA.SD^2 / GAMMA.MEAN
      
    #aN  <- round( N.sample * data.in$pop.wt[row], 0 )
      aN  <- N.sample
    
    for(aSIM in 1:CI.sample){ #
      
      select   <- max(select) + 1:aN
      
      # assign sociodemographic variables:
      vSEX[select]      <- as.character(gamma.dat$sex)
      vAGE[select]      <- as.character(gamma.dat$age)
      vSES[select]      <- as.character(gamma.dat$ses)
      vSIM[select]      <- aSIM
      
      # assign drawn APC and CD vars
      vAPC.BASE[select]             <- APC.draw[aSIM]
      vAPC.BASE.DRINKER[select]     <- APC.DRINKER.draw[aSIM]
      
      # assign LA / CD status
      vLA.ref[select]                       <- as.logical(rbinom(n = aN, size = 1, prob = LA.draw[aSIM]))
      vCD.ref[select][vLA.ref[select] == F] <- as.logical(rbinom(n = sum(vLA.ref[select] == F), size = 1, prob = ( CD.draw[aSIM]/(1-LA.draw[aSIM]) ) ))
      
      # draw drinking levels and adjust so mean matches GAMMA.MEAN (mean drinking level among drinkers)
      vDL.ref[select][vCD.ref[select]==T]   <- round( rgamma(n = sum(vCD.ref[select]==T), shape = KAPPA[aSIM], scale = THETA[aSIM]), 3)
      vDL.ref[select][vCD.ref[select]==T]   <- vDL.ref[select][vCD.ref[select]==T] * 
        (GAMMA.MEAN[aSIM] / mean(vDL.ref[select][vCD.ref[select]==T]) )
      vDL.ref[select][vCD.ref[select]==F]   <- NA
      
      # draw HED status by drinking level risk status:
      
        # low risk:
        HED.select <- vDL.ref[select][vCD.ref[select]==T] <= risk.threshold.1[gamma.dat$sex]
        
        if(HED.draw.risk.low[aSIM] == 1){
          vHED.ref[select][vCD.ref[select]==T][HED.select] <- T
        } else {
          vHED.ref[select][vCD.ref[select]==T][HED.select] <- rbinom(n = sum(HED.select), size = 1, prob = HED.draw.risk.low[aSIM])
          }

        
        # mid risk:
        HED.select <- vDL.ref[select][vCD.ref[select]==T] > risk.threshold.1[gamma.dat$sex] & vDL.ref[select][vCD.ref[select]==T] <= risk.threshold.2[gamma.dat$sex]
        
        if(HED.draw.risk.mid[aSIM] == 1){
          vHED.ref[select][vCD.ref[select]==T][HED.select] <- T
        } else {
          vHED.ref[select][vCD.ref[select]==T][HED.select] <- rbinom(n = sum(HED.select), size = 1, prob = HED.draw.risk.mid[aSIM])
        }
        
        
        # high risk:
        HED.select <- vDL.ref[select][vCD.ref[select]==T] > risk.threshold.2[gamma.dat$sex]
        
        if(HED.draw.risk.high[aSIM] == 1){
          vHED.ref[select][vCD.ref[select]==T][HED.select] <- T
        } else {
          vHED.ref[select][vCD.ref[select]==T][HED.select] <- rbinom(n = sum(HED.select), size = 1, prob = HED.draw.risk.high[aSIM])
        }
      
    }
      
  }
  end <- max(select)
  rm(select, gamma.dat)
  
  dat <- data.table(id = 1:end, sex = vSEX[1:end], age = vAGE[1:end], ses = vSES[1:end], sim = vSIM[1:end], 
                    LA.y0 = vLA.ref[1:end], CD.y0 = vCD.ref[1:end], 
                    DL.y0 = vDL.ref[1:end], HED.y0 = as.logical(vHED.ref[1:end]))
  dat[, FD.y0 := ifelse(LA.y0 == F & CD.y0 == F, T, F)]
  dat[, HED.y0 := ifelse(DL.y0 >= 60, T, HED.y0)]
  dat[, HED.y0 := ifelse(CD.y0 == F, NA, HED.y0)]
  dat[, age := factor(age, levels = age.groups, ordered = T)]
  
  # check:
  dat[sex == "female" & age == "15-34" & ses == "mid", table(LA.y0, CD.y0, useNA = "always")]
  dat[sex == "female" & age == "15-34" & ses == "mid", table(LA.y0, FD.y0, useNA = "always")]
  dat[sex == "female" & age == "15-34" & ses == "high", table(CD.y0, DL.y0 > 0, useNA = "always")]
  dat[, mean(CD.y0), by = .(sex,age,ses)]
  
  # mean LA / FD / CD
  dat[, .(mean(LA.y0),mean(FD.y0),mean(CD.y0)), by = .(sex,age,ses)][,.(V1 + V2 + V3)]
  
  dat$attenuate.year.count <- NA_integer_
  
  #  C) Loop for each year:
  #  ...................................
  
  # Prepare attenuation parameters
  aAttenuate.threshold <- aAttenuate[attenuation < 1][,min(year)]
  aAttenuate[, att.factor := -c(NA, diff(attenuation, 1))]
  aAttenuate[is.na(att.factor) & year == 1, att.factor := ifelse(attenuation <0 , attenuation, 0)]
  
  #y <- 1
  years <- length(years.lab)
  for(y in 1:(years)){ #
    
    print(paste0("Year ", years.lab[y], ": Simulating data for ", unique(aScenario[year == y]$coverage)*100, "% coverage"))
    
    # D1) Prepare 
    # ----------------------------
    
    # ALC VARIABLES OF THIS YEAR **.work = drinking level to work with for current year
    # => use values from last year
    
    dat$LA.work <- dat[, .SD, .SDcols = names(dat) %like% paste0("LA.y",y-1)]
    dat$FD.work <- dat[, .SD, .SDcols = names(dat) %like% paste0("FD.y",y-1)]
    dat$CD.work <- dat[, .SD, .SDcols = names(dat) %like% paste0("CD.y",y-1)]
    dat$DL.work <- dat[, .SD, .SDcols = names(dat) %like% paste0("DL.y",y-1)]
    dat$HED.work <- dat[, .SD, .SDcols = names(dat) %like% paste0("HED.y",y-1)]
    
    # add variable indicating consecutive year
    dat$temp <- y
    names(dat)[names(dat) == "temp"] <- paste0("year",y)
    
    
    # D2) Attenuate effects from previous years
    # ----------------------------
    
    ##  NOTE: THE CODE RESETS THE LAG STRUCTURE OF THE EFFECTS EVERY TIME A NEW INTERVENTION WAS GIVEN !!
    # e.g. intervention at t0, 50% at t1; new intervention at t1, again 50% of new effect at t2
    
    if(y >= 2){
      
      # get temporary data with DL / HED to be changed:
      temp <- copy(dat[!is.na(attenuate.year.count), .SD, .SDcols = names(dat) %like% "attenuate|DL.y|HED.y"])
      names(temp)[names(temp) == paste0("DL.y",y-1)]   <- "DLlag1"
      names(temp)[names(temp) == paste0("DL.y",y-2)]   <- "DLlag2"
      names(temp)[names(temp) == paste0("DL.y",y-3)]   <- "DLlag3"
      names(temp)[names(temp) == paste0("DL.y",y-4)]   <- "DLlag4"
      names(temp)[names(temp) == paste0("DL.y",y-5)]   <- "DLlag5"
      names(temp)[names(temp) == paste0("DL.y",y-6)]   <- "DLlag6"
      names(temp)[names(temp) == paste0("DL.y",y-7)]   <- "DLlag7"
      names(temp)[names(temp) == paste0("DL.y",y-8)]   <- "DLlag8"
      names(temp)[names(temp) == paste0("DL.y",y-9)]   <- "DLlag9"
      names(temp)[names(temp) == paste0("DL.y",y-10)]  <- "DLlag10"
      
      names(temp)[names(temp) == paste0("HED.y",y-1)]   <- "HEDlag1"
      names(temp)[names(temp) == paste0("HED.y",y-2)]   <- "HEDlag2"
      names(temp)[names(temp) == paste0("HED.y",y-3)]   <- "HEDlag3"
      names(temp)[names(temp) == paste0("HED.y",y-4)]   <- "HEDlag4"
      names(temp)[names(temp) == paste0("HED.y",y-5)]   <- "HEDlag5"
      names(temp)[names(temp) == paste0("HED.y",y-6)]   <- "HEDlag6"
      names(temp)[names(temp) == paste0("HED.y",y-7)]   <- "HEDlag7"
      names(temp)[names(temp) == paste0("HED.y",y-8)]   <- "HEDlag8"
      names(temp)[names(temp) == paste0("HED.y",y-9)]   <- "HEDlag9"
      names(temp)[names(temp) == paste0("HED.y",y-10)]  <- "HEDlag10"
      
      # attenuate based on specified attenuation parameters:
      temp$dl.corrected <- NA_real_
      temp$HED.corrected <- as.logical(NA_integer_)
      temp$DL.interv.effect  <- NA_real_
      temp$HED.interv.effect  <- NA_real_
      
      # no attenuation:
      temp[attenuate.year.count < aAttenuate.threshold, dl.corrected := DLlag1]
      temp[attenuate.year.count < aAttenuate.threshold, HED.corrected := HEDlag1]
      
      # with attenuation:
      # => take last drinking level which was not attenuated + add effects of attenuation
      temp[attenuate.year.count >= aAttenuate.threshold]
      
      for (ayc in unique(temp[attenuate.year.count >= aAttenuate.threshold]$attenuate.year.count)){
        
        # for DL: calculate intervention effect
        temp[attenuate.year.count == ayc]$DL.interv.effect <- 
          
          temp[attenuate.year.count == ayc, .SD, .SDcols = names(temp) %like% paste0("DLlag",ayc+1) ]  -
          temp[attenuate.year.count == ayc, .SD, .SDcols = names(temp) %like% paste0("DLlag",ayc) ] 
        
        # add attenuated effect
        temp[attenuate.year.count == ayc, dl.corrected := DLlag1 + DL.interv.effect * aAttenuate[year == ayc]$att.factor ]
        
        # for HED: identify individuals who changed from HED to non-HED
        temp[attenuate.year.count == ayc]$HED.interv.effect <- 
          
          temp[attenuate.year.count == ayc, .SD, .SDcols = names(temp) %like% paste0("HEDlag",ayc+1) ]!=
          temp[attenuate.year.count == ayc, .SD, .SDcols = names(temp) %like% paste0("HEDlag",ayc) ] 
        
        # recalculate likelihood to relapse to HED
        # if at least 2 years have passed since successful intervention, relapse to HED at 50% chance (assumption)
        temp[attenuate.year.count == ayc, HED.corrected := ifelse(HED.interv.effect == F, HEDlag1, 
                                                                  rbinom(n = .N, size = 1, prob = 0.5))]
        
      }
      
      temp[is.na(dl.corrected)] # should be none
      temp[is.na(HED.corrected)] # should be none
      
      temp[!is.na(attenuate.year.count), ]
      
      # replace old with corrected DL:
      dat[!is.na(attenuate.year.count), DL.work := round( temp$dl.corrected , 3) ]
      
      # replace old with corrected HED:
      dat[!is.na(attenuate.year.count), HED.work := temp$HED.corrected ]
    }
    
    
    # C3) Account for APC time trend
    # ----------------------------
    if(y >= 2){
      
    # for each sex-age-ses group: calculate change in APC
      APC.adjust <- merge(data.in[year == years.lab[y-1]][,.(sex,age,ses,APC_y0 = APC)],
                          data.in[year == years.lab[y]][,.(sex,age,ses,APC_y1 = APC)], by = c("sex","age","ses"), all.x = T)
      APC.adjust[, APC.diff := APC_y1 - APC_y0]
      APC.adjust[, apc.change := APC.diff / APC_y0]
      
      dat <- merge(dat, APC.adjust[,.(sex,age,ses,apc.change)], by = c("sex","age","ses"), all.x = T)
      dat[, DL.work := DL.work * (1 + apc.change) ] 
      dat$apc.change <- NULL
      rm(APC.adjust)
    
    }
    
    # C4) Account for LA / CD / HED time trend
    # ----------------------------
    
    if(y >= 2){
      
      print(paste0("Accounting for changes in patterns of alcohol use between year ",years.lab[y-1]," and ",years.lab[y]))
      
      # calculate change rate between previous and current year
        sub <- copy(data.in[year %in% years.lab[c(y,y-1)],.(year,sex,age,ses,LA,FD = 1-LA-CD,CD,HED = HED_DRINKER)]) #,CD.SE
        sub <- dcast(sub, sex + age + ses ~ year, value.var = c("LA","FD","CD","HED")) #,"CD.SE"
        names(sub)[names(sub) %like% years.lab[y-1]] <- gsub(years.lab[y-1], 1, names(sub)[names(sub) %like% years.lab[y-1]])
        names(sub)[names(sub) %like% years.lab[y]] <- gsub(years.lab[y], 2, names(sub)[names(sub) %like% years.lab[y]])
        sub[,  ':=' (LA_diff = LA_2 - LA_1, CD_diff = CD_2 - CD_1, HED_diff = HED_2 - HED_1) ]
        sub[,  ':=' (LA_change = ifelse(LA_diff > 0, "more LA", ifelse(LA_diff < 0, "less LA", "no change")),
                     CD_change = ifelse(CD_diff > 0, "more CD", ifelse(CD_diff < 0, "less CD", "no change")),
                     HED_change = ifelse(HED_diff > 0, "more HED", ifelse(HED_diff < 0, "less HED", "no change")))]
        sub[ LA_change == "more LA", LA_change.more := LA_diff / (FD_1) ] # take new LA cases from FD ???
        sub[ LA_change == "less LA", LA_change.less := abs(LA_diff / (LA_1)) ] # all former LA cases to be taken from LA only (and assigned to CD)
        sub[ CD_change == "more CD", CD_change.more.fromFD := CD_diff*0.5 / (FD_1) ] # all new CD cases to be taken from FD only
        sub[ CD_change == "more CD", CD_change.more.fromLA := CD_diff*0.5 / (LA_1) ] # all new CD cases to be taken from FD only
        sub[ CD_change == "less CD", CD_change.less := abs(CD_diff / (CD_1)) ] # all former CD cases to be taken from FD only (and assigned to FD)
        sub[ HED_change== "more HED",HED_change.more:= HED_diff / (CD_1 - HED_1)] # take new HED cases from non-heavy drinking CD
        sub[ HED_change== "less HED",HED_change.less:= abs(HED_diff / HED_1)] # remove HED cases from heavy drinking CD
      
      # apply change rate to sample
        dat <- merge(dat, sub[,.(sex,age,ses,
                                 LA_change,LA_change.less,LA_change.more,
                                 CD_change,CD_change.less,CD_change.more.fromFD,CD_change.more.fromLA,
                                 HED_change,HED_change.less,HED_change.more)], by = c("sex","age","ses"), all.x = T)
        rm(sub)
        
        # if "more LA" => take new LA from FD
        dat[LA_change == "more LA" & FD.work == T, new.LA := as.logical( rbinom(.N, size = 1, prob = ifelse(LA_change.more>1,1,LA_change.more) ) ), 
            by = .(sex,age,ses)]
        dat[new.LA == T, ':=' (LA.work = T, FD.work = F)]
        
        # if "less LA" => move LA to CD
        dat[LA_change == "less LA" & LA.work == T, new.CD := as.logical( rbinom(.N, size = 1, prob = ifelse(LA_change.less>1,1,LA_change.less) ) ), 
            by = .(sex,age,ses)]
        dat[new.CD == T, ':=' (LA.work = F, CD.work = T)]
        
        # if "more CD" => take new CD from both FD and LA (50/50)
        dat[CD_change == "more CD" & FD.work == T, new.CD := as.logical( rbinom(.N, size = 1, prob = ifelse(CD_change.more.fromFD>1,1,CD_change.more.fromFD) ) ), 
            by = .(sex,age,ses)]
        dat[CD_change == "more CD" & LA.work == T, new.CD := as.logical( rbinom(.N, size = 1, prob = ifelse(CD_change.more.fromLA>1,1,CD_change.more.fromLA)) ), 
            by = .(sex,age,ses)]
        dat[new.CD == T, ':=' (CD.work = T, FD.work = F, LA.work = F)]
        
        # if "less CD" => move CD to FD
        dat[CD_change == "less CD" & CD.work == T, new.FD := as.logical( rbinom(.N, size = 1, prob = ifelse(CD_change.less>1,1,CD_change.less) ) ), 
            by = .(sex,age,ses)]
        dat[new.FD == T, ':=' (CD.work = F, FD.work = T)]
        
        # if "more HED" => take new HED from non-heavy drinking CD
        dat[HED_change == "more HED" & CD.work == T & HED.work == F, new.HED := as.logical( rbinom(.N, size = 1, prob = ifelse(HED_change.more>1,1,HED_change.more) ) ), 
            by = .(sex,age,ses)]
        dat[new.HED == T, ':=' (HED.work = T)]
      
        # if "less HED" => remove HED from heavy drinking CD to non-heavy drinking CD
        dat[HED_change == "less HED" & CD.work == T & HED.work == T & DL.work < 60, new.non_HED := as.logical( rbinom(.N, size = 1, prob = ifelse(HED_change.less>1,1,HED_change.less) ) ), 
            by = .(sex,age,ses)]
        dat[new.non_HED == T, ':=' (HED.work = F)]
        
      # get DL and HED for new drinkers from current year
      dat[new.CD == T,.(sex,age,ses,CD.work,DL.work,HED.work)]
      
      draw.dat <- copy( data.in[year == years.lab[y],.(sex,age,ses,APC,CD,HED.LOW,HED.MID,HED.HIGH)] )
      
      draw.dat[, ':=' (GAMMA.MEAN = (APC / CD) * 1000 * 0.789 / 365,
                       GAMMA.SD   = ifelse(sex == "male", 1.171 * GAMMA.MEAN , 1.258 * GAMMA.MEAN))]
      draw.dat[, ':=' (KAPPA = GAMMA.MEAN^2 / GAMMA.SD^2, 
                       THETA = GAMMA.SD^2 / GAMMA.MEAN)]
      
      #int <- unique(interaction(dat[,.(sex,age,ses)]))[1]
      for(int in unique(interaction(dat[,.(sex,age,ses)]))){ # loop over all sex/age/ses groups and draw drinking levels / HED status
        
        # drinking levels
        dat[new.CD == T & interaction(sex,age,ses) == int, DL.work := round( rgamma(n = .N, 
                                                                                    shape = draw.dat[interaction(sex,age,ses) == int]$KAPPA,
                                                                                    scale = draw.dat[interaction(sex,age,ses) == int]$THETA), 3)]
        
        # HED for low risk drinkers:
        dat[new.CD == T & interaction(sex,age,ses) == int & DL.work <= risk.threshold.1[sex],.(sex,age,ses,DL.work,HED.work)]
        dat[new.CD == T & interaction(sex,age,ses) == int & DL.work <= risk.threshold.1[sex], 
            HED.work := rbinom(n = .N, size = 1, prob = draw.dat[interaction(sex,age,ses) == int]$HED.LOW)]
        
        # HED for mid risk drinkers:
        dat[new.CD == T & interaction(sex,age,ses) == int & DL.work > risk.threshold.1[sex] & DL.work <= risk.threshold.2[sex],.(sex,age,ses,DL.work,HED.work)]
        dat[new.CD == T & interaction(sex,age,ses) == int & DL.work > risk.threshold.1[sex] & DL.work <= risk.threshold.2[sex],
            HED.work := rbinom(n = .N, size = 1, prob = draw.dat[interaction(sex,age,ses) == int]$HED.MID)]
        
        # HED for high risk drinkers:
        dat[new.CD == T & interaction(sex,age,ses) == int & DL.work > risk.threshold.2[sex] ,.(sex,age,ses,DL.work,HED.work)]
        dat[new.CD == T & interaction(sex,age,ses) == int & DL.work > risk.threshold.2[sex],
            HED.work := rbinom(n = .N, size = 1, prob = draw.dat[interaction(sex,age,ses) == int]$HED.HIGH)]
        dat[, HED.work := ifelse(DL.work >= 60, T, HED.work)]
        
      } # end of new DL / HED value loop
      
      # remove vars:
      dat <- dat[,.SD,.SDcols = names(dat)[!names(dat) %like% "LA_change|CD_change|HED_change|^new"]]
      rm(draw.dat)
      
    } #end of LA / CD / HED changes
    
    
    # C5) SBIRT impact
    # ----------------------------
    
    # GET RISK CATEGORY & PHC PARAMETERS:
    
        # DL category
        dat$RISK <- NA_character_
        dat[sex == "female" , RISK :=  ifelse(DL.work <= risk.threshold.1["female"], "low",
                                              ifelse(DL.work <= risk.threshold.2["female"], "mid", "high")) ]
        dat[sex == "male" , RISK :=  ifelse(DL.work <= risk.threshold.1["male"], "low",
                                            ifelse(DL.work <= risk.threshold.2["male"], "mid", "high")) ]
        dat[, RISK := factor(RISK, levels = c("low","mid","high"), ordered = T)]
        
        # PHC visit probability
        dat <- merge(dat, data.in[year == years.lab[y],.(sex,age,ses,
                                                         PHC.risk.low,PHC.risk.low.SE,
                                                         PHC.risk.mid,PHC.risk.mid.SE,
                                                         PHC.risk.high,PHC.risk.high.SE)], by = c("sex","age","ses"))
        
        dat$PHC.prob <- NA_real_
        dat[RISK == "low", PHC.prob := rnorm(n = .N, mean = PHC.risk.low, sd = PHC.risk.low.SE), by = .(sex,age,ses)]
        dat[RISK == "mid", PHC.prob := rnorm(n = .N, mean = PHC.risk.mid, sd = PHC.risk.mid.SE), by = .(sex,age,ses)]
        dat[RISK == "high", PHC.prob := rnorm(n = .N, mean = PHC.risk.high, sd = PHC.risk.high.SE), by = .(sex,age,ses)]
        dat[, PHC.prob := ifelse(PHC.prob < 0, 0, ifelse(PHC.prob > 1, 1, PHC.prob))]
        
        dat$PHC.risk.low <- dat$PHC.risk.mid <- dat$PHC.risk.high <- NULL
        dat$PHC.risk.low.SE <- dat$PHC.risk.mid.SE <- dat$PHC.risk.high.SE <- NULL
        
        # select persons based on PHC admission probability, SBIRT probability
        dat[DL.work > 0, PHC.adm := as.logical(rbinom(.N, size = 1, prob = PHC.prob)) ]
        
    # GET SBIRT PARAMETERS:
        
        ##  screening
        dat[, screened := ifelse(PHC.adm == T,as.logical(rbinom(n = .N, size = 1, prob = aScenario[year == y]$coverage)),F) ]
        
        ##  brief intervention for RISK == low
            dat$BI.receive.prob <- NA_real_
            dat[screened == T & RISK == "mid"]$BI.receive.prob <- aProb.intervention
            dat[!is.na(BI.receive.prob), BI.received := as.logical(rbinom(.N, size = 1, prob = BI.receive.prob)) ]
            dat[is.na(BI.received), BI.received := F]
            
            ### draw BI effects for DL based on proportional reduction
            dat$BI.effect.DL <- NA_real_
            dat[sex == "female" & BI.received == T, BI.effect.DL := rnorm(mean = as.numeric(aEff.bi.dl[var %like% "est",.SD, .SDcols = "female"]),
                                                                          sd = as.numeric(aEff.bi.dl[var %like% "sd",.SD, .SDcols = "female"]),
                                                                          n = .N) ]
            dat[sex == "male" & BI.received == T, BI.effect.DL := rnorm(mean = as.numeric(aEff.bi.dl[var %like% "est",.SD, .SDcols = "male"]), 
                                                                        sd = as.numeric(aEff.bi.dl[var %like% "sd",.SD, .SDcols = "male"]), 
                                                                        n = .N) ]
            
            ### draw BI effects for HED based on risk reduction
            
            # first determine current HED risk
              dat[CD.work == T, HED_current.risk := mean(HED.work), by = .(sex,age,ses,sim,RISK)]
            # then determine new (reduced) risk, drawn from parameter distribution:
              dat[, HED_new.risk := HED_current.risk + rnorm(1, mean = aEff.bi.hed["est"], aEff.bi.hed["sd"]), by = .(sex,age,ses,sim,RISK)]
              dat[HED_new.risk<0, HED_new.risk := 0]
            # 1 - the ratio between new and current risk is the probability for which current HED may remit
              dat[, BI.effect.HED.prob := 1 - (HED_new.risk / HED_current.risk)]
              dat[BI.effect.HED.prob < 0, BI.effect.HED.prob := 0]
              
              dat$BI.effect.HED <- logical(nrow(dat))
              dat[BI.received == T & HED_current.risk > 0, BI.effect.HED := as.logical(rbinom(n = .N,
                                                                                              size = 1,
                                                                                              prob = BI.effect.HED.prob)), by = .(sex,age,ses,sim,RISK)]
              
              dat$HED_current.risk <- dat$HED_new.risk <- dat$BI.effect.HED.prob <- NULL
        
        ##  referral to treatment
            dat$RT.receive.prob <- NA_real_
            dat[screened == T & RISK == "high"]$RT.receive.prob <- aProb.intervention
            dat[!is.na(RT.receive.prob), RT.received := as.logical(rbinom(.N, size = 1, prob = RT.receive.prob)) ]
            dat[is.na(RT.received), RT.received := F]
            
            ### draw RT effects for DL based on proportional reduction
            dat$RT.effect.DL <- NA_real_
            dat[sex == "female" & RT.received == T, RT.effect.DL := rnorm(mean = as.numeric(aEff.rt.dl[var %like% "est",.SD, .SDcols = "female"]), 
                                                                          sd = as.numeric(aEff.rt.dl[var %like% "sd",.SD, .SDcols = "female"]), 
                                                                          n = .N) ]
            dat[sex == "male" & RT.received == T, RT.effect.DL := rnorm(mean = as.numeric(aEff.rt.dl[var %like% "est",.SD, .SDcols = "male"]),
                                                                        sd = as.numeric(aEff.rt.dl[var %like% "sd",.SD, .SDcols = "male"]), 
                                                                        n = .N) ]
        
            ### draw RT effects for HED based on risk reduction
            
            # first determine current HED risk
            dat[CD.work == T, HED_current.risk := mean(HED.work), by = .(sex,age,ses,sim,RISK)]
            # then determine new (reduced) risk, drawn from parameter distribution:
            dat[, HED_new.risk := HED_current.risk + rnorm(1, mean = aEff.rt.hed["est"], aEff.rt.hed["sd"]), by = .(sex,age,ses,sim,RISK)]
            dat[HED_new.risk<0, HED_new.risk := 0]
            # the ratio between new and current risk is the probability for which current HED may remit
            dat[, RT.effect.HED.prob := 1 - (HED_new.risk / HED_current.risk)]
            dat[RT.effect.HED.prob < 0, RT.effect.HED.prob := 0]
            
            dat$RT.effect.HED <- logical(nrow(dat))
            dat[RT.received == T & HED_current.risk > 0, RT.effect.HED := as.logical(rbinom(n = .N,
                                                                     size = 1,
                                                                     prob = RT.effect.HED.prob)) ]
            
            dat$HED_current.risk <- dat$HED_new.risk <- dat$RT.effect.HED.prob <- NULL
            
            
    #  APPLY BI / RT effects => NEW DL / CD / FD / HED
            
        dat[screened == T, DL.new := round(ifelse(BI.received == T, DL.work * (1+BI.effect.DL),
                                                  ifelse(RT.received == T, DL.work * (1+RT.effect.DL), DL.work )),3) ]
        dat[, DL.new := round(ifelse(is.na(DL.new), DL.work , DL.new ),3) ]
        
        # change drinking status if DL.new < 0
        dat[DL.new < 0, ':=' (FD.new = T, CD.new = F)]
        dat[, ':=' (FD.new = ifelse(is.na(FD.new), FD.work , FD.new ),
                    CD.new = ifelse(is.na(CD.new), CD.work , CD.new )) ]
        dat[, DL.new := round(ifelse(DL.new < 0, 0 , DL.new ),3) ] # correct if DL falls below 0
        
        ##  get new HED (only possible if DL < 60)
        dat[screened == T & DL.new < 60, HED.new := ifelse(BI.received == T & BI.effect.HED == T, F,
                                                           ifelse(RT.received == T & RT.effect.HED == T, F, HED.work )) ] # recode HED to FALSE if BI/RT effect = T
        dat[, HED.new := ifelse(is.na(HED.new), HED.work, HED.new)]
        
    
    #  ATTENUATE YEAR COUNT
    
      ##  if drinking level changed in this round, set "attenuation" variable to 1, otherwise + 1
      dat[, attenuate.year.count := ifelse( BI.received == T | RT.received == T, 1, 
                                            ifelse(attenuate.year.count >= 1, attenuate.year.count + 1, NA)) ]
      
      # remove and rename year-specific parameters
      dat <- copy(dat[, .SD, .SDcols = !names(dat) %like% "prob|effect"])
      
      
      names(dat)[names(dat) %like% "RISK$|PHC.adm$|screened$|BI.received$|RT.received$"] <- 
        paste0(names(dat)[names(dat) %like% "RISK$|PHC.adm$|screened$|BI.received$|RT.received$"],".y",y)
      
      names(dat)[names(dat) %like% "LA.work"] <- paste0("LA.y",y)
      names(dat)[names(dat) %like% "FD.new"] <- paste0("FD.y",y)
      names(dat)[names(dat) %like% "CD.new"] <- paste0("CD.y",y)
      names(dat)[names(dat) %like% "DL.new"] <- paste0("DL.y",y)
      names(dat)[names(dat) %like% "HED.new"] <- paste0("HED.y",y)
      
      gc()
      
      # add year-specific intervention variable:
      dat$interv <- NA
      dat[ , interv := as.logical(
        rowSums( dat[, .SD, .SDcols = names(dat) %like% paste0("BI.received.y",y,"|RT.received.y",y)], na.rm = T ) > 0) ]
      dat[ , interv := ifelse(is.na(interv), F, interv) ]  
      names(dat)[names(dat) == "interv"]  <- paste0("interv.y", y)
    
    
  } # END OF YEAR LOOP
  
  # get baseline categorization of drinking levels:
  dat[sex == "female" , RISK.BL :=  ifelse(DL.y0 <= risk.threshold.1["female"], "low",
                                           ifelse(DL.y0 <= risk.threshold.2["female"], "mid", "high")) ]
  
  dat[sex == "male" , RISK.BL :=  ifelse(DL.y0 <= risk.threshold.1["male"], "low",
                                         ifelse(DL.y0 <= risk.threshold.2["male"], "mid", "high")) ]
  dat[, RISK.BL := factor(RISK.BL, levels = c("low","mid","high"), ordered = T)]
  
  # make SES a factor
  dat[, ses := factor(ses, levels = c("low","mid","high"), ordered = T)]
  
  ##  RESHAPE
  
  print(" ... RESHAPING AND CLEANING DATA ... ")
  
  # rename year variables:
  
  for(y in 1:years){
    
    names(dat)[names(dat) %like% paste0("y",y,"$")] <- gsub(paste0("y",y), paste0("y",years.lab[y]), names(dat)[names(dat) %like% paste0("y",y,"$")] )  
    
  }
  
  
  dat.ext <- dat[,.SD, .SDcols = names(dat) %like% "LA|FD|CD|HED|id|sex|age|ses|sim|DL.y[0-9]|HED.y[0-9]|RISK.y[1-9]|interv.y[1-9]|screened.y[1-9]|RISK.BL" &
                   !names(dat) %like% "work"]
  
  # extract drinking levels, HED, screening and intervention variable
  
    # drinking statuses:
    add1 <- melt(dat.ext[,.SD, .SDcols = names(dat.ext) %like% "^id|sex|ses|age|sim|LA.y[0-9]|FD.y[0-9]|CD.y[0-9]|HED.y[0-9]"],
                 id.vars = c("id","sex","age","ses","sim","LA.y0","FD.y0","CD.y0","HED.y0"), value.name = c("LA","FD","CD","HED"), variable.name = "year", 
                 measure.vars = patterns("LA.y2","FD.y2","CD.y2","HED.y2"))
    
    
    # drinking level, baseline risk, screened & interv
    add2 <- melt(dat.ext[,.SD, .SDcols = names(dat.ext) %like% "^id|sex|ses|age|sim|DL.y[0-9]|RISK.y[0-9]|RISK.BL|screened|interv"],
                 id.vars = c("id","sex","age","ses","sim","DL.y0","RISK.BL"), value.name = c("drink.level","RISK","screened","interv"), variable.name = "year",
                 patterns("DL.y2","RISK.y2","screened","interv"))
    
  print(" ... ... ... ... ")
  gc()
  
    # years
    add1$year <- as.numeric(add1$year) + years.lab[1] -1
    add2$year <- as.numeric(add2$year) + years.lab[1] -1
  
  print(" ... ... ... ... ")
  
    dat.all <- merge(add1, add2, by = c("sim", "id", "year", "sex", "age", "ses"))
    
    gc()
    rm(add1); rm(add2)
    gc()
  
  
  ##  Add scenario, coverage and population weight:
  
    dat.all$scenario <- unique(aScenario$scenario)
    dat.all <- merge(dat.all, aScenario[,.(year = year + min(years.lab)-1,coverage)], by = c("year"), all.x = T)
    
    dat.all <- merge(dat.all, data.in[,.(year,sex,age,ses = as.character(ses),pop)], by = c("year","sex","age","ses"), all.x = T)
    
    dat.all[!complete.cases(dat.all[,.(year,sex,age,ses,sim,id,LA,FD,CD)])] # should be none
    
  ##  Add screening and intervention variable:
    
    dat.all[, screened.any.year := any(screened == T), by = .(scenario, sim, id, sex, age, ses)]
    dat.all[, interv.any.year := any(interv == T), by = .(scenario, sim, id, sex, age, ses)]
    
  ##  RETURN
    
  print("  DONE :) ")
  
  return(dat.all[,.SD, by = .(scenario, sim, id, sex, age, ses, DL.y0, RISK.BL)])
  
  
} # end of function
```
