## Supplementary material for "Can alcohol consumption in Germany be reduced by alcohol screening, brief intervention and referral to treatment in primary health care? Results of a simulation study": S1 File: SBIRT impact_4_sensitivity analyses.R

```
# ========================================================================================================================================================================================================
# ========================================================================================================================================================================================================
# ========================================================================================================================================================================================================

#	CODE FOR MANUSCRIPT: 
# Can alcohol consumption in Germany be reduced by alcohol screening, brief intervention and referral to treatment in primary health care? Results of a simulation study

# Author of code:
# Jakob Manthey

# ========================================================================================================================================================================================================
# ========================================================================================================================================================================================================
# ========================================================================================================================================================================================================
# ========================================================================================================================================================================================================
# ========================================================================================================================================================================================================
# ========================================================================================================================================================================================================

# _____________________________________________________________________
#	LOAD ALL NECESSARY LIBRARIES:
# _____________________________________________________________________

# ................................................................................
#	LIBRARIES
# ................................................................................

library (data.table)
library (Hmisc)
library (openxlsx)
library (MASS)
library (stringr)
library (dglm)

# ========================================================================================================================================================================================================
# ========================================================================================================================================================================================================
# ========================================================================================================================================================================================================
# ========================================================================================================================================================================================================
# ========================================================================================================================================================================================================
# ========================================================================================================================================================================================================


# ========================================================================================================================================================================================================
# ========================================================================================================================================================================================================
# ========================================================================================================================================================================================================
# ========================================================================================================================================================================================================
# ========================================================================================================================================================================================================
# ========================================================================================================================================================================================================

    
#	___________________________________________
#	COMBINE VECTORS INTO DATA TABLE
#	___________________________________________

##  Sensitivity analyses
      sens0 <- FUN_SIM_(data.in = base.data, N.sample = 100, CI.sample = 100, aScenario = main.analyses[scenario == 0], aSeed = 46483, aAttenuate = attenuation.sens) #2.9%
      sens1 <- FUN_SIM_(data.in = base.data, N.sample = 100, CI.sample = 100, aScenario = main.analyses[scenario == 1], aSeed = 46483, aAttenuate = attenuation.sens) #0%
      sens2 <- FUN_SIM_(data.in = base.data, N.sample = 100, CI.sample = 100, aScenario = main.analyses[scenario == 2], aSeed = 46483, aAttenuate = attenuation.sens) #10%
      sens3 <- FUN_SIM_(data.in = base.data, N.sample = 100, CI.sample = 100, aScenario = main.analyses[scenario == 3], aSeed = 46483, aAttenuate = attenuation.sens) #25%
      sens4 <- FUN_SIM_(data.in = base.data, N.sample = 100, CI.sample = 100, aScenario = main.analyses[scenario == 4], aSeed = 46483, aAttenuate = attenuation.sens) #50%
      sens5 <- FUN_SIM_(data.in = base.data, N.sample = 100, CI.sample = 100, aScenario = main.analyses[scenario == 5], aSeed = 46483, aAttenuate = attenuation.sens) #75%
      
      
      sens.dat <- rbindlist(mget(ls()[ls() %like% "^sens[0-9]"]))      
      
      
#	==========================================================================================================================================
#	==========================================================================================================================================
#	==========================================================================================================================================
#	==========================================================================================================================================
#	==========================================================================================================================================
#	==========================================================================================================================================

#	___________________________________________
# CALIBRATION OF ESTIMATES
#	___________________________________________

##  RATIONALE: POINT ESTIMATES OF BASELINE SCENARIO (0) NEED TO BE EXACT THE SAME AS INPUT DATA.
##  ==> FOR EACH SUBGROUP, DETERMINE CALIBRATION FACTOR
      
#  a) aggregate simulated data at point estimates
sens.modelled <-  sens.dat[scenario == 0, .(LA.mod = mean(LA), 
                                           FD.mod = mean(FD), 
                                           CD.mod = mean(CD),
                                           HED.mod = mean(HED, na.rm = T), 
                                           DL.mod = mean(drink.level,na.rm = T)),
                          by = .(sex, age, ses, year)]
      
#  b) combine with input data
sens.calib <-  merge(sens.modelled,
                     alc.data[,.(sex, age, ses, year,
                                 LA.in = LA, FD.in = 1-LA-CD, CD.in = CD, HED.in = HED_DRINKER, 
                                 DL.in = (APC / CD) * 1000 * 0.789 / 365)], 
                     by = c("year","sex","age","ses"), all = T)

#  c) get calibration factors:
sens.calib <- sens.calib[, .( year,sex,age,ses,
                              LA.calib = LA.in / LA.mod,
                              FD.calib = FD.in / FD.mod,
                              CD.calib = CD.in / CD.mod,
                              HED.calib = HED.in / HED.mod,
                              DL.calib = DL.in / DL.mod)]

# d) remove objects
rm(sens.modelled)

#	___________________________________________
# AGGREGATION OF ESTIMATES
#	___________________________________________

##  LEVEL 0: ESTIMATES PER SIM
#   .........................................

#  a) for each simulation
agg.sens0 <- sens.dat[, .(LA = mean(LA), 
                      FD = mean(FD), 
                      CD = mean(CD),
                      HED = mean(HED, na.rm = T), 
                      DL = mean(drink.level,na.rm = T), 
                      RISK.LOW = mean(RISK == "low",na.rm = T), 
                      RISK.MID = mean(RISK == "mid",na.rm = T), 
                      RISK.HIGH = mean(RISK == "high",na.rm = T), 
                      screened = mean(screened, na.rm = T),
                      screened.any.year = mean(screened.any.year, na.rm = T),
                      interv = mean(interv, na.rm = T),
                      interv.any.year = mean(interv.any.year, na.rm = T),
                      pop = unique(pop)),
                   by = .(scenario, coverage, sex, age, ses, year, sim)]

# b) correct estimates with calibration factors:
agg.sens0 <- merge(agg.sens0, sens.calib, by = c("year","sex","age","ses"), all.x = T)
agg.sens0[, ':=' (LA = LA * LA.calib,
                FD = FD * FD.calib,
                CD = CD * CD.calib,
                HED = HED * HED.calib,
                DL = DL * DL.calib)]


# c) normalize LA+CD+FD to 1

  # [only for non-baseline scenarios]

agg.sens0[, ':=' (normalize = LA + FD + CD)]
agg.sens0[, ':=' (LA = LA / normalize,
               FD = FD / normalize,
               CD = CD / normalize)]
agg.sens0$normalize <- NULL

# d) get APC
agg.sens0[, APC := (DL * CD) / (1000 * 0.789 / 365)]

# e) clean
agg.sens0$LA.calib <- agg.sens0$FD.calib <- agg.sens0$CD.calib <- agg.sens0$HED.calib <- agg.sens0$DL.calib <- NULL
rm(sens.calib)

##  AGGREGATE LEVEL 1: ALL SUBGROUPS
#   .........................................

# a) get PE and CI
agg.sens1 <- agg.sens0[,.(LA = quantile(LA, 0.5), LA.lower = quantile(LA, 0.025), LA.upper = quantile(LA, 0.975),
                    FD = quantile(FD, 0.5), FD.lower = quantile(FD, 0.025), FD.upper = quantile(FD, 0.975),
                    CD = quantile(CD, 0.5), CD.lower = quantile(CD, 0.025), CD.upper = quantile(CD, 0.975),
                    HED = quantile(HED, 0.5), HED.lower = quantile(HED, 0.025), HED.upper = quantile(HED, 0.975),
                    DL = quantile(DL, 0.5), DL.lower = quantile(DL, 0.025), DL.upper = quantile(DL, 0.975),
                    RISK.LOW = quantile(RISK.LOW, 0.5), RISK.LOW.lower = quantile(RISK.LOW, 0.025), RISK.LOW.upper = quantile(RISK.LOW, 0.975),
                    RISK.MID = quantile(RISK.MID, 0.5), RISK.MID.lower = quantile(RISK.MID, 0.025), RISK.MID.upper = quantile(RISK.MID, 0.975),
                    RISK.HIGH = quantile(RISK.HIGH, 0.5), RISK.HIGH.lower = quantile(RISK.HIGH, 0.025), RISK.HIGH.upper = quantile(RISK.HIGH, 0.975),
                    APC = quantile(APC, 0.5), APC.lower = quantile(APC, 0.025), APC.upper = quantile(APC, 0.975),
                    screened = quantile(screened, 0.5), screened.lower = quantile(screened, 0.025), screened.upper = quantile(screened, 0.975),
                    screened.any.year = quantile(screened.any.year, 0.5), screened.any.year.lower = quantile(screened.any.year, 0.025), screened.any.year.upper = quantile(screened.any.year, 0.975),
                    interv = quantile(interv, 0.5), interv.lower = quantile(interv, 0.025), interv.upper = quantile(interv, 0.975),
                    interv.any.year = quantile(interv.any.year, 0.5), interv.any.year.lower = quantile(interv.any.year, 0.025), interv.any.year.upper = quantile(interv.any.year, 0.975)
                    ),by = .(scenario, coverage, sex, age, ses, year, pop)]

# b) compare estimated and input data:
      
      figure.yes <- F
      
      if(figure.yes == T){
        
        pdat <- merge(agg.sens1[scenario == 0,.(sex, age, ses, year, 
                                             LA.mod = LA, FD.mod = FD, CD.mod = CD, HED.mod = HED, DL.mod = DL, APC.mod = APC)],
                      alc.data[,.(year, sex, age, ses, LA.in = LA, FD.in = 1-LA-CD, CD.in = CD, HED.in = HED_DRINKER, DL.in = (APC / CD) * 1000 * 0.789 / 365, APC.in = APC )], 
                      by = c("year","sex","age","ses"), all.x = T)
        
        pdat <- melt(pdat, value.name = c("LA", "FD", "CD", "HED", "DL", "APC"), measure.vars = patterns("LA.", "FD.", "CD.", "HED.", "DL.", "APC"), variable.name = "source")
        pdat <- melt(pdat, id.vars = c("year","sex","age","ses", "source"), variable.name = "drink.var")
        pdat[, source := ifelse(source == 1, "model", "input")]
        
        pdf("0_sens_drinking status manipulations_l1.pdf", height = 15, width = 10)
        
        ggplot(pdat[sex == "female" & !drink.var %like% "DL|APC"], aes(x = year, y = value, color = source)) + 
          ggtitle("female") +
          theme_minimal() + facet_grid(age + ses ~ drink.var, scales = "free") + 
          geom_line()
        
        ggplot(pdat[sex == "male"  & !drink.var %like% "DL|APC"], aes(x = year, y = value, color = source)) + 
          ggtitle("male") +
          theme_minimal() + facet_grid(age + ses ~ drink.var, scales = "free") + 
          geom_line()
        
        ggplot(pdat[drink.var %like% "DL"], aes(x = year, y = value, color = source)) + 
          ggtitle("DL") +
          theme_minimal() + facet_grid(age + ses ~ sex, scales = "free") + 
          geom_line()
        
        ggplot(pdat[drink.var %like% "APC"], aes(x = year, y = value, color = source)) + 
          ggtitle("APC") +
          theme_minimal() + facet_grid(age + ses ~ sex, scales = "free") + 
          geom_line()
        
        dev.off() # should be all on one line!
        
      }


##  LEVEL 2: GROUPS
#   .........................................

#   this needs two steps: first weighted mean within sim, second quantiles across all sims

agg.sens0[sex == "female" & year == 2009 & scenario == 1, wtd.mean(LA, pop)]

# a) BY SEX
agg.sens2a <- agg.sens0[,.(age = "15-99", ses = "all", 
                     LA = wtd.mean(LA,pop), 
                     FD = wtd.mean(FD,pop), 
                     CD = wtd.mean(CD,pop),
                     HED = wtd.mean(HED,pop), 
                     DL = wtd.mean(DL,pop),
                     RISK.LOW = wtd.mean(RISK.LOW,pop), 
                     RISK.MID = wtd.mean(RISK.MID,pop), 
                     RISK.HIGH = wtd.mean(RISK.HIGH,pop), 
                     APC = wtd.mean(APC,pop),
                     screened = wtd.mean(screened,pop),
                     screened.any.year = wtd.mean(screened.any.year,pop),
                     interv = wtd.mean(interv,pop),
                     interv.any.year = wtd.mean(interv.any.year,pop),
                     pop = sum(pop)),
                  by = .(scenario, coverage, sex, year, sim)]

agg.sens2a <- agg.sens2a[,.(LA = quantile(LA, 0.5), LA.lower = quantile(LA, 0.025), LA.upper = quantile(LA, 0.975),
                      FD = quantile(FD, 0.5), FD.lower = quantile(FD, 0.025), FD.upper = quantile(FD, 0.975),
                      CD = quantile(CD, 0.5), CD.lower = quantile(CD, 0.025), CD.upper = quantile(CD, 0.975),
                      HED = quantile(HED, 0.5), HED.lower = quantile(HED, 0.025), HED.upper = quantile(HED, 0.975),
                      DL = quantile(DL, 0.5), DL.lower = quantile(DL, 0.025), DL.upper = quantile(DL, 0.975),
                      RISK.LOW = quantile(RISK.LOW, 0.5), RISK.LOW.lower = quantile(RISK.LOW, 0.025), RISK.LOW.upper = quantile(RISK.LOW, 0.975),
                      RISK.MID = quantile(RISK.MID, 0.5), RISK.MID.lower = quantile(RISK.MID, 0.025), RISK.MID.upper = quantile(RISK.MID, 0.975),
                      RISK.HIGH = quantile(RISK.HIGH, 0.5), RISK.HIGH.lower = quantile(RISK.HIGH, 0.025), RISK.HIGH.upper = quantile(RISK.HIGH, 0.975),
                      APC = quantile(APC, 0.5), APC.lower = quantile(APC, 0.025), APC.upper = quantile(APC, 0.975),
                      screened = quantile(screened, 0.5), screened.lower = quantile(screened, 0.025), screened.upper = quantile(screened, 0.975),
                      screened.any.year = quantile(screened.any.year, 0.5), screened.any.year.lower = quantile(screened.any.year, 0.025), screened.any.year.upper = quantile(screened.any.year, 0.975),
                      interv = quantile(interv, 0.5), interv.lower = quantile(interv, 0.025), interv.upper = quantile(interv, 0.975),
                      interv.any.year = quantile(interv.any.year, 0.5), interv.any.year.lower = quantile(interv.any.year, 0.025), interv.any.year.upper = quantile(interv.any.year, 0.975)
                      ),
                    by = .(scenario, coverage, sex, age, ses, year, pop)]
  
# b) BY AGE
agg.sens2b <- agg.sens0[, .(sex = "total", ses = "all", 
                      LA = wtd.mean(LA,pop), 
                      FD = wtd.mean(FD,pop), 
                      CD = wtd.mean(CD,pop),
                      HED = wtd.mean(HED,pop), 
                      DL = wtd.mean(DL,pop),
                      RISK.LOW = wtd.mean(RISK.LOW,pop), 
                      RISK.MID = wtd.mean(RISK.MID,pop), 
                      RISK.HIGH = wtd.mean(RISK.HIGH,pop), 
                      APC = wtd.mean(APC,pop),
                      screened = wtd.mean(screened,pop),
                      screened.any.year = wtd.mean(screened.any.year,pop),
                      interv = wtd.mean(interv,pop),
                      interv.any.year = wtd.mean(interv.any.year,pop),
                      pop = sum(pop)),
                  by = .(scenario, coverage, age, year, sim)]

agg.sens2b <- agg.sens2b[,.(LA = quantile(LA, 0.5), LA.lower = quantile(LA, 0.025), LA.upper = quantile(LA, 0.975),
                      FD = quantile(FD, 0.5), FD.lower = quantile(FD, 0.025), FD.upper = quantile(FD, 0.975),
                      CD = quantile(CD, 0.5), CD.lower = quantile(CD, 0.025), CD.upper = quantile(CD, 0.975),
                      HED = quantile(HED, 0.5), HED.lower = quantile(HED, 0.025), HED.upper = quantile(HED, 0.975),
                      DL = quantile(DL, 0.5), DL.lower = quantile(DL, 0.025), DL.upper = quantile(DL, 0.975),
                      RISK.LOW = quantile(RISK.LOW, 0.5), RISK.LOW.lower = quantile(RISK.LOW, 0.025), RISK.LOW.upper = quantile(RISK.LOW, 0.975),
                      RISK.MID = quantile(RISK.MID, 0.5), RISK.MID.lower = quantile(RISK.MID, 0.025), RISK.MID.upper = quantile(RISK.MID, 0.975),
                      RISK.HIGH = quantile(RISK.HIGH, 0.5), RISK.HIGH.lower = quantile(RISK.HIGH, 0.025), RISK.HIGH.upper = quantile(RISK.HIGH, 0.975),
                      APC = quantile(APC, 0.5), APC.lower = quantile(APC, 0.025), APC.upper = quantile(APC, 0.975),
                      screened = quantile(screened, 0.5), screened.lower = quantile(screened, 0.025), screened.upper = quantile(screened, 0.975),
                      screened.any.year = quantile(screened.any.year, 0.5), screened.any.year.lower = quantile(screened.any.year, 0.025), screened.any.year.upper = quantile(screened.any.year, 0.975),
                      interv = quantile(interv, 0.5), interv.lower = quantile(interv, 0.025), interv.upper = quantile(interv, 0.975),
                      interv.any.year = quantile(interv.any.year, 0.5), interv.any.year.lower = quantile(interv.any.year, 0.025), interv.any.year.upper = quantile(interv.any.year, 0.975)
                      ),
                   by = .(scenario, coverage, sex, age, ses, year, pop)]

# c) BY SES
agg.sens2c <- agg.sens0[, .(sex = "total", age = "15-99", 
                      LA = wtd.mean(LA,pop), 
                      FD = wtd.mean(FD,pop), 
                      CD = wtd.mean(CD,pop),
                      HED = wtd.mean(HED,pop), 
                      DL = wtd.mean(DL,pop),
                      RISK.LOW = wtd.mean(RISK.LOW,pop), 
                      RISK.MID = wtd.mean(RISK.MID,pop), 
                      RISK.HIGH = wtd.mean(RISK.HIGH,pop), 
                      APC = wtd.mean(APC,pop),
                      screened = wtd.mean(screened,pop),
                      screened.any.year = wtd.mean(screened.any.year,pop),
                      interv = wtd.mean(interv,pop),
                      interv.any.year = wtd.mean(interv.any.year,pop),
                      pop = sum(pop)),
                  by = .(scenario, coverage, ses, year, sim)]

agg.sens2c <- agg.sens2c[,.(LA = quantile(LA, 0.5), LA.lower = quantile(LA, 0.025), LA.upper = quantile(LA, 0.975),
                      FD = quantile(FD, 0.5), FD.lower = quantile(FD, 0.025), FD.upper = quantile(FD, 0.975),
                      CD = quantile(CD, 0.5), CD.lower = quantile(CD, 0.025), CD.upper = quantile(CD, 0.975),
                      HED = quantile(HED, 0.5), HED.lower = quantile(HED, 0.025), HED.upper = quantile(HED, 0.975),
                      DL = quantile(DL, 0.5), DL.lower = quantile(DL, 0.025), DL.upper = quantile(DL, 0.975),
                      RISK.LOW = quantile(RISK.LOW, 0.5), RISK.LOW.lower = quantile(RISK.LOW, 0.025), RISK.LOW.upper = quantile(RISK.LOW, 0.975),
                      RISK.MID = quantile(RISK.MID, 0.5), RISK.MID.lower = quantile(RISK.MID, 0.025), RISK.MID.upper = quantile(RISK.MID, 0.975),
                      RISK.HIGH = quantile(RISK.HIGH, 0.5), RISK.HIGH.lower = quantile(RISK.HIGH, 0.025), RISK.HIGH.upper = quantile(RISK.HIGH, 0.975),
                      APC = quantile(APC, 0.5), APC.lower = quantile(APC, 0.025), APC.upper = quantile(APC, 0.975),
                      screened = quantile(screened, 0.5), screened.lower = quantile(screened, 0.025), screened.upper = quantile(screened, 0.975),
                      screened.any.year = quantile(screened.any.year, 0.5), screened.any.year.lower = quantile(screened.any.year, 0.025), screened.any.year.upper = quantile(screened.any.year, 0.975),
                      interv = quantile(interv, 0.5), interv.lower = quantile(interv, 0.025), interv.upper = quantile(interv, 0.975),
                      interv.any.year = quantile(interv.any.year, 0.5), interv.any.year.lower = quantile(interv.any.year, 0.025), interv.any.year.upper = quantile(interv.any.year, 0.975)
                      ),
                   by = .(scenario, coverage, sex, age, ses, year, pop)]

# d) compare estimated and input data:
    
    if(figure.yes == T){

        #   by sex (as example)
        pdat <- merge(agg.sens2a[scenario == 0,.(sex, age, ses, year, 
                                             LA.mod = LA, FD.mod = FD, CD.mod = CD, HED.mod = HED, DL.mod = DL, APC.mod = APC)],
                      alc.data[,.(age = "15-99", ses = "all", LA.in = wtd.mean(LA,pop), FD.in = wtd.mean(1-LA-CD, pop), CD.in = wtd.mean(CD, pop), HED.in = wtd.mean(HED_DRINKER, pop), DL.in = wtd.mean((APC / CD) * 1000 * 0.789 / 365,pop), APC.in = wtd.mean(APC,pop) ), by = .(year,sex)], 
                      by = c("year","sex","age","ses"), all.x = T)
        
        pdat <- melt(pdat, value.name = c("LA", "FD", "CD", "HED", "DL", "APC"), measure.vars = patterns("LA.", "FD.", "CD.", "HED.", "DL.", "APC"), variable.name = "source")
        pdat <- melt(pdat, id.vars = c("year","sex","age","ses", "source"), variable.name = "drink.var")
        pdat[, source := ifelse(source == 1, "model", "input")]
        
        pdf("0_sens_drinking status manipulations_l2.pdf", height = 5, width = 6)
        
        ggplot(pdat[!drink.var %like% "DL|APC"], aes(x = year, y = value, color = source)) + 
          ggtitle("drinking statuses") +
          theme_minimal() + facet_grid(sex ~ drink.var, scales = "free") + 
          geom_line()
        
        ggplot(pdat[drink.var %like% "DL"], aes(x = year, y = value, color = source)) + 
          ggtitle("DL") +
          theme_minimal() + facet_grid(sex ~ ., scales = "free") + 
          geom_line()
        
        ggplot(pdat[drink.var %like% "APC"], aes(x = year, y = value, color = source)) + 
          ggtitle("APC") +
          theme_minimal() + facet_grid(sex ~ ., scales = "free") + 
          geom_line()
        
        dev.off() # should be all on one line!
    
    }


##  LEVEL 3: TOTAL
#   .........................................

#   this needs two steps: first weighted mean within sim, second quantiles across all sims

agg.sens3 <- agg.sens0[, .(sex = "total", age = "15-99", ses = "all", 
                     LA = wtd.mean(LA,pop), 
                     FD = wtd.mean(FD,pop), 
                     CD = wtd.mean(CD,pop),
                     HED = wtd.mean(HED,pop), 
                     DL = wtd.mean(DL,pop),
                     RISK.LOW = wtd.mean(RISK.LOW,pop), 
                     RISK.MID = wtd.mean(RISK.MID,pop), 
                     RISK.HIGH = wtd.mean(RISK.HIGH,pop), 
                     APC = wtd.mean(APC,pop),
                     screened = wtd.mean(screened,pop),
                     screened.any.year = wtd.mean(screened.any.year,pop),
                     interv = wtd.mean(interv,pop),
                     interv.any.year = wtd.mean(interv.any.year,pop),
                     pop = sum(pop)),
                  by = .(scenario, coverage, year, sim)]

agg.sens3 <- agg.sens3[,.(LA = quantile(LA, 0.5), LA.lower = quantile(LA, 0.025), LA.upper = quantile(LA, 0.975),
                    FD = quantile(FD, 0.5), FD.lower = quantile(FD, 0.025), FD.upper = quantile(FD, 0.975),
                    CD = quantile(CD, 0.5), CD.lower = quantile(CD, 0.025), CD.upper = quantile(CD, 0.975),
                    HED = quantile(HED, 0.5), HED.lower = quantile(HED, 0.025), HED.upper = quantile(HED, 0.975),
                    DL = quantile(DL, 0.5), DL.lower = quantile(DL, 0.025), DL.upper = quantile(DL, 0.975),
                    RISK.LOW = quantile(RISK.LOW, 0.5), RISK.LOW.lower = quantile(RISK.LOW, 0.025), RISK.LOW.upper = quantile(RISK.LOW, 0.975),
                    RISK.MID = quantile(RISK.MID, 0.5), RISK.MID.lower = quantile(RISK.MID, 0.025), RISK.MID.upper = quantile(RISK.MID, 0.975),
                    RISK.HIGH = quantile(RISK.HIGH, 0.5), RISK.HIGH.lower = quantile(RISK.HIGH, 0.025), RISK.HIGH.upper = quantile(RISK.HIGH, 0.975),
                    APC = quantile(APC, 0.5), APC.lower = quantile(APC, 0.025), APC.upper = quantile(APC, 0.975),
                    screened = quantile(screened, 0.5), screened.lower = quantile(screened, 0.025), screened.upper = quantile(screened, 0.975),
                    screened.any.year = quantile(screened.any.year, 0.5), screened.any.year.lower = quantile(screened.any.year, 0.025), screened.any.year.upper = quantile(screened.any.year, 0.975),
                    interv = quantile(interv, 0.5), interv.lower = quantile(interv, 0.025), interv.upper = quantile(interv, 0.975),
                    interv.any.year = quantile(interv.any.year, 0.5), interv.any.year.lower = quantile(interv.any.year, 0.025), interv.any.year.upper = quantile(interv.any.year, 0.975)
                    ),
                   by = .(scenario, coverage, sex, age, ses, year, pop)]

# b) compare estimated and input data:

    if(figure.yes == T){
  
        pdat <- merge(agg.sens3[scenario == 0,.(sex, age, ses, year, 
                                             LA.mod = LA, FD.mod = FD, CD.mod = CD, HED.mod = HED, DL.mod = DL, APC.mod = APC)],
                      alc.data[,.(sex = "total", age = "15-99", ses = "all", LA.in = wtd.mean(LA,pop), FD.in = wtd.mean(1-LA-CD, pop), CD.in = wtd.mean(CD, pop), HED.in = wtd.mean(HED_DRINKER, pop), DL.in = wtd.mean((APC / CD) * 1000 * 0.789 / 365,pop), APC.in = wtd.mean(APC,pop) ), by = .(year)], 
                      by = c("year","sex","age","ses"), all.x = T)
        
        pdat <- melt(pdat, value.name = c("LA", "FD", "CD", "HED", "DL", "APC"), measure.vars = patterns("LA.", "FD.", "CD.", "HED.", "DL.", "APC"), variable.name = "source")
        pdat <- melt(pdat, id.vars = c("year","sex","age","ses", "source"), variable.name = "drink.var")
        pdat[, source := ifelse(source == 1, "model", "input")]
        
        pdf("0_sens_drinking status manipulations_l3.pdf", height = 5, width = 6)
        
        ggplot(pdat[!drink.var %like% "DL|APC"], aes(x = year, y = value, color = source)) + 
          ggtitle("drinking statuses") +
          theme_minimal() + facet_grid(. ~ drink.var, scales = "free") + 
          geom_line()
        
        ggplot(pdat[drink.var %like% "DL"], aes(x = year, y = value, color = source)) + 
          ggtitle("DL") +
          theme_minimal() + #facet_grid(sex ~ ., scales = "free") + 
          geom_line()
        
        ggplot(pdat[drink.var %like% "APC"], aes(x = year, y = value, color = source)) + 
          ggtitle("APC") +
          theme_minimal() + #facet_grid(sex ~ ., scales = "free") + 
          geom_line()
        
        dev.off() # should be all on one line!

    }

##  COMBINE ALL LEVELS TO ONE SINGLE DATA TABLE
#   .........................................
    
    sens.data <- rbind(agg.sens1, agg.sens2a, agg.sens2b, agg.sens2c, agg.sens3)
    
    sens.data[, scenario.lab := ifelse(scenario == 0, "baseline: 2.9%", 
                                  ifelse(scenario == 1, "0%",
                                         ifelse(scenario == 2, "10%", 
                                                ifelse(scenario == 3, "25%",
                                                       ifelse(scenario == 4, "50%", 
                                                              ifelse(scenario == 5, "75%", ""))))))]
    sens.data[, ses := factor(ses, levels = c("all","low","mid","high"), ordered = T)]
    factor(sens.data$ses)
    
    
    
            
# ========================================================================================================================================================================================================
# ========================================================================================================================================================================================================
# ========================================================================================================================================================================================================
# ========================================================================================================================================================================================================
# ========================================================================================================================================================================================================
# ========================================================================================================================================================================================================

# ................................................................................
#	FUNCTION FOR CALCULATING THE DIFFERENCE BETWEEN MAIN AND SENSITIVITY ANALYSES AT END OF TEN YEAR PERIOD
# ................................................................................

add1 <- copy(agg.l0)
add1$source <- "main"
add2 <- copy(agg.sens0)
add2$source <- "sens"

main.and.sens <- rbind(add1, add2)
    
FUN_DIFF_SENS_ <- function(input = main.and.sens, 
                           aComparator = 1,
                           out.format = "table",
                           cross.aggregate = T
){
  
  ##  PREPARE
  
  var.select <- names(input)[names(input) %like% paste0("^scenario|^id|^sim|^year|^sex|^age|^ses|^pop|^DL$|^source$")]
  comp.dat <- input[year %in% c(max(year)) & scenario %in% c(aComparator), .SD, .SDcols = var.select ][order(year)]
  rm(var.select)
  
  ##  determine difference for output variables DL
  comp.dat.diff <- dcast(comp.dat[,.(sim,sex,age,ses,pop,source,DL)], formula = sim + sex + age + ses + pop ~ source, value.var = "DL")
  comp.dat.diff[, DL := (sens - main)/main]
  
  ##  aggregate differences
  
  if(cross.aggregate == T){
    
    ##  BY SEX
    comp.dat.diff.sex <- comp.dat.diff[,.(age = "15-99", ses = "all", 
                                          DL = wtd.mean(DL,pop),
                                          pop = sum(pop)),
                                       by = .(sex, sim)]
    
    comp.dat.diff.sex <- comp.dat.diff.sex[,.(DL = quantile(DL, 0.5), DL.lower = quantile(DL, 0.025), DL.upper = quantile(DL, 0.975)),
                                           by = .(sex, age, ses)]
    
    ##  BY AGE
    comp.dat.diff.age <- comp.dat.diff[,.(sex = "total", ses = "all", 
                                          DL = wtd.mean(DL,pop),
                                          pop = sum(pop)),
                                       by = .(age, sim)]
    
    comp.dat.diff.age <- comp.dat.diff.age[,.(DL = quantile(DL, 0.5), DL.lower = quantile(DL, 0.025), DL.upper = quantile(DL, 0.975)),
                                           by = .(sex, age, ses)]
    
    ##  BY SES
    comp.dat.diff.ses <- comp.dat.diff[,.(sex = "total", age = "15-99", 
                                          DL = wtd.mean(DL,pop),
                                          pop = sum(pop)),
                                       by = .(ses, sim)]
    
    comp.dat.diff.ses <- comp.dat.diff.ses[,.(DL = quantile(DL, 0.5), DL.lower = quantile(DL, 0.025), DL.upper = quantile(DL, 0.975)),
                                           by = .(sex, age, ses)]
    
    ##  BY ALL
    comp.dat.diff.all <- comp.dat.diff[,.(sex = "total", age = "15-99", ses = "all", 
                                          DL = wtd.mean(DL,pop),
                                          pop = sum(pop)),
                                       by = .(sim)]
    
    comp.dat.diff.all <- comp.dat.diff.all[,.(DL = quantile(DL, 0.5), DL.lower = quantile(DL, 0.025), DL.upper = quantile(DL, 0.975)),
                                           by = .(sex, age, ses)]
    
    ##  COMBINE
    
    comp.out <- rbind(comp.dat.diff.sex, comp.dat.diff.age, comp.dat.diff.ses, comp.dat.diff.all)
    
    
  } else {
    
    ##  BY SEX, AGE, SES
    comp.out <- comp.dat.diff[,.(DL = wtd.mean(DL,pop),
                                 pop = sum(pop)),
                              by = .(sex, age, ses, sim)]
    
    comp.out <- comp.out[,.(DL = quantile(DL, 0.5), DL.lower = quantile(DL, 0.025), DL.upper = quantile(DL, 0.975)),
                         by = .(sex, age, ses)]
    
  }
  
  ##  FORMAT    
  
  comp.out$comparator <- aComparator
  
  if(out.format == "table"){
    
    var.select <- c("DL","DL.lower","DL.upper")
    
    comp.out[, (var.select) := lapply(.SD, function(x){x * 100}), .SDcols = var.select]
    
    comp.out[, (var.select) := lapply(.SD, format, digits = 0, nsmall = 1), .SDcols = var.select]
    
    comp.out[, DL.out := paste0(DL, "% (",DL.lower," to ", DL.upper,"%)")]
    
    rm(var.select)
    
    return(comp.out[,.(sex,age,comparator,ses,DL = DL.out)])
    
  } else {
    
    return(comp.out)  
    
  }
  
}
```
