## Supplementary material for "Can alcohol consumption in Germany be reduced by alcohol screening, brief intervention and referral to treatment in primary health care? Results of a simulation study": S1 File: SBIRT impact_5_results.R

```
# ========================================================================================================================================================================================================
# ========================================================================================================================================================================================================
# ========================================================================================================================================================================================================

#	CODE FOR MANUSCRIPT: 
# Can alcohol consumption in Germany be reduced by alcohol screening, brief intervention and referral to treatment in primary health care? Results of a simulation study

# Author of code:
# Jakob Manthey

# ========================================================================================================================================================================================================
# ========================================================================================================================================================================================================
# ========================================================================================================================================================================================================
# ========================================================================================================================================================================================================
# ========================================================================================================================================================================================================
# ========================================================================================================================================================================================================

rm(list=ls())
DATE  <- 210408

# _____________________________________________________________________
#	RUN SIMULATION
# _____________________________________________________________________

##  1) source function
# ................................................................................

source(paste0('C:/Data.work/Publications/PlosOne_SBIRT Germany/SBIRT impact_1_prepare data.R'))
source(paste0('C:/Data.work/Publications/PlosOne_SBIRT Germany/SBIRT impact_2_simulation.R'))
source(paste0('C:/Data.work/Publications/PlosOne_SBIRT Germany/SBIRT impact_3_main analyses.R'))
source(paste0('C:/Data.work/Publications/PlosOne_SBIRT Germany/SBIRT impact_4_sensitivity analyses.R'))

# ========================================================================================================================================================================================================
# ========================================================================================================================================================================================================
# ========================================================================================================================================================================================================
# ========================================================================================================================================================================================================
# ========================================================================================================================================================================================================
# ========================================================================================================================================================================================================

# _____________________________________________________________________
#	REPORTING IN "RESULTS" SECTION
# _____________________________________________________________________

##  1) Baseline scenario
# ................................................................................

# APC & CD
data[scenario == 0 & sex == "total" & age == "15-99" & ses == "all" & year %in% c(2009,2018), .(year, APC, APC.lower, APC.upper)]
data[scenario == 0 & sex == "total" & age == "15-99" & ses == "all" & year %in% c(2009,2018), .(year, CD, CD.lower, CD.upper)]
data[scenario == 0 & sex == "total" & age == "15-99" & ses == "low" & year %in% c(2009,2018), .(year, CD, CD.lower, CD.upper)]
data[scenario == 0 & sex == "total" & age == "15-99" & ses == "high" & year %in% c(2009,2018), .(year, CD, CD.lower, CD.upper)]

# RISK
data[sex == "total" & age == "15-99" & ses == "all" & year %in% c(2009,2018) & scenario == 0, .(year, RISK.LOW, RISK.LOW.lower, RISK.LOW.upper)]
data[sex == "total" & age == "15-99" & ses == "all" & year %in% c(2009,2018) & scenario == 0, .(year, RISK.MID, RISK.MID.lower, RISK.MID.upper)]
data[sex == "total" & age == "15-99" & ses == "all" & year %in% c(2009,2018) & scenario == 0, .(year, RISK.HIGH, RISK.HIGH.lower, RISK.HIGH.upper)]

# HED
data[sex == "total" & age == "15-99" & ses == "all" & year %in% c(2009,2018) & scenario == 0, .(year, HED, HED.lower, HED.upper)]

##  2) Coverage of SBIRT
# ................................................................................

data[sex == "total" & age == "15-99" & ses == "all" & year == 2018, .(coverage, screened.any.year, screened.any.year.lower, screened.any.year.upper)]
data[sex == "total" & age == "15-99" & ses == "all" & year == 2018, .(coverage, interv.any.year, interv.any.year.lower, interv.any.year.upper)]

sim.dat[year == 2009 & scenario == 0, mean(screened, na.rm = T)]
sim.dat[year == 2009 & scenario == 0, mean(interv, na.rm = T)]
sim.dat[year == 2013 & scenario == 0, mean(interv.any.year, na.rm = T)]
sim.dat[year == 2013 & scenario == 0, mean(interv.any.year, na.rm = T)]

sim.dat[year == 2013 & scenario == 3 & interv == T, table(RISK)]

#   % screened
data[scenario == 2 & CAT == "all" & sex == "total" & age == "25-49" & year == 10, .(setting, coverage, screen.mean)]
agg.all[scenario == 7 & CAT == "all" & sex == "total" & age == "25-49" & year == 10, .(setting, coverage, screen.mean)]

##  3) Subgroup differences
# ................................................................................

diff.scen1 <- FUN_DIFF_(aComparator = 1, cross.aggregate = F, out.format = "other")
diff.scen2 <- FUN_DIFF_(aComparator = 2, cross.aggregate = F, out.format = "other")
diff.scen3 <- FUN_DIFF_(aComparator = 3, cross.aggregate = F, out.format = "other")
diff.scen4 <- FUN_DIFF_(aComparator = 4, cross.aggregate = F, out.format = "other")
diff.scen5 <- FUN_DIFF_(aComparator = 5, cross.aggregate = F, out.format = "other")

diff <- rbind(diff.scen1, diff.scen2, diff.scen3, diff.scen4, diff.scen5)[,.(comparator, sex, age, ses, DL, DL.lower, DL.upper)]

#   for Figure 4 - females:
diff[sex == "female" & comparator == 1 & DL.upper < 0,] # none
diff[sex == "female" & comparator == 2 & DL.upper < 0,] # none
diff[sex == "female" & comparator == 3 & DL.upper < 0,] # 1
diff[sex == "female" & comparator == 4 & DL.upper < 0,] # 5
diff[sex == "female" & comparator == 5 & DL.upper < 0,] # 6

#   for Figure 5 - males:
diff[sex == "male" & comparator == 1 & DL.upper < 0,] # none
diff[sex == "male" & comparator == 2 & DL.upper < 0,] # none
diff[sex == "male" & comparator == 3 & DL.upper < 0,] # 5
diff[sex == "male" & comparator == 4 & DL.upper < 0,] # 10
diff[sex == "male" & comparator == 4 & DL.upper > 0,] # 2
diff[sex == "male" & comparator == 5 & DL.upper < 0,] # 11

##  4) Sensitivity analyses
# ................................................................................

##  run comparisons
diff.sens.scen1 <- FUN_DIFF_SENS_(aComparator = 1)
diff.sens.scen2 <- FUN_DIFF_SENS_(aComparator = 2)
diff.sens.scen3 <- FUN_DIFF_SENS_(aComparator = 3)
diff.sens.scen4 <- FUN_DIFF_SENS_(aComparator = 4)
diff.sens.scen5 <- FUN_DIFF_SENS_(aComparator = 5)

diff.sens.dat    <- rbind(diff.sens.scen1, diff.sens.scen2, diff.sens.scen3, diff.sens.scen4, diff.sens.scen5)

# scenario 1
data[sex == "total" & age == "15-99" & ses == "all" & scenario == 1 & year == 2018,.(APC)]
sens.data[sex == "total" & age == "15-99" & ses == "all" & scenario == 1 & year == 2018,.(APC)]
diff.sens.dat[sex == "total" & age == "15-99" & ses == "all" & comparator == 1]

# scenario 2
data[sex == "total" & age == "15-99" & ses == "all" & scenario == 2 & year == 2018,.(APC)]
sens.data[sex == "total" & age == "15-99" & ses == "all" & scenario == 2 & year == 2018,.(APC)]
diff.sens.dat[sex == "total" & age == "15-99" & ses == "all" & comparator == 2]

# scenario 3
data[sex == "total" & age == "15-99" & ses == "all" & scenario == 3 & year == 2018,.(APC)]
sens.data[sex == "total" & age == "15-99" & ses == "all" & scenario == 3 & year == 2018,.(APC)]
diff.sens.dat[sex == "total" & age == "15-99" & ses == "all" & comparator == 3]

# scenario 4
data[sex == "total" & age == "15-99" & ses == "all" & scenario == 4 & year == 2018,.(APC)]
sens.data[sex == "total" & age == "15-99" & ses == "all" & scenario == 4 & year == 2018,.(APC)]
diff.sens.dat[sex == "total" & age == "15-99" & ses == "all" & comparator == 4]

# scenario 5
data[sex == "total" & age == "15-99" & ses == "all" & scenario == 5 & year == 2018,.(APC)]
sens.data[sex == "total" & age == "15-99" & ses == "all" & scenario == 5 & year == 2018,.(APC)]
diff.sens.dat[sex == "total" & age == "15-99" & ses == "all" & comparator == 5]


#	==========================================================================================================================================
#	==========================================================================================================================================
#	==========================================================================================================================================
#	==========================================================================================================================================
#	==========================================================================================================================================
#	==========================================================================================================================================

##  reset WD:    
setwd( paste0("C:/Data.work/Publications/PlosOne_SBIRT Germany/output/",DATE) )

#	___________________________________________
#	FIGURES
#	___________________________________________

#colors.3.lila <- c( '#8c96c6','#8856a7','#810f7c' )
#colors.5.lila <- c( '#edf8fb','#b3cde3','#8c96c6','#8856a7','#810f7c')
colors.3.green <- rev(c('#a6611a' , '#dfc27d' , '#80cdc1'))
#colors.3 <- c("#F7EE7F", "#F1A66A", "#F26157")

c8Panel_c <- c('#785144', '#814959', '#5E589A', '#5D7CA3', '#62AAAB', '#72B78A', '#CCCD94', '#E0B7C5')

##  2) Individual drinking trajectories
#   .........................................

# baseline with 1 intervention 
id.select <- sim.dat[scenario == 0 & DL.y0 %between% c(30,50) & sex == "female" & age == "35-49" & ses == "mid" 
                     & year == 2010 & interv == T,.(choose = sum(interv)), by = .(id,sex) ][choose == 1][1]$id
pdat <- copy( sim.dat[id == id.select & scenario == 1] )

# no intervention
id.select <- sim.dat[scenario == 1 & DL.y0 %between% c(30,50) & sex == "female" & age == "35-49" & ses == "mid"]$id[45]
pdat <- rbind( pdat, copy( sim.dat[id == id.select & scenario == 0] ) )

# 3 interventions
id.select <- sim.dat[scenario == 5 & DL.y0 %between% c(30,50) & sex == "female" & age == "35-49" & ses == "mid"
                     & (year != 2011),.(choose = sum(interv)), by = .(id,sex,sim) ][choose == 3][1]$id
pdat <- rbind( pdat, copy( sim.dat[id == id.select & scenario == 5] ) )

pdat <- pdat[,.(id = as.factor(id),sex,year,base = DL.y0, drink.level,screened,interv)][order(id,year)]

ggplot(pdat, aes(year,drink.level,color = id)) +
  theme_bw() + 
  geom_vline(xintercept = pdat[interv == T & id == unique(pdat$id)[1]]$year, color = c8Panel_c[1], linetype = 1, size = 1) + 
  geom_vline(xintercept = pdat[interv == T & id == unique(pdat$id)[2]]$year, color = c8Panel_c[4], linetype = 1, size = 1) +
  geom_vline(xintercept = pdat[interv == T & id == unique(pdat$id)[3]]$year, color = c8Panel_c[6], linetype = 1, size = 1) + 
  geom_point(size = 4) + 
  geom_line(size = 2) + 
  scale_color_manual(values = c8Panel_c[c(1,4,6)]) +
  guides(color = FALSE) + 
  scale_x_continuous(name = "Year", breaks = c(2009:2018)) + 
  scale_y_continuous(name = "Drinking level (g pure alcohol per day)") 

ggsave("Figure 2_individual drinking trajectory.png", width = 8)
ggsave("Fig 2.tiff", width = 8, dpi = 300, unit = "in")

##  3) Variation of screening and intervention rates by scenario
#   .........................................

pdat1  <- data[sex == "total" & age == "15-99" & ses == "all" & year == 2018,.(coverage, var = "screening", est = screened.any.year, low = screened.any.year.lower, upp = screened.any.year.upper)]
pdat2  <- data[sex == "total" & age == "15-99" & ses == "all" & year == 2018,.(coverage, var = "intervention", est = interv.any.year, low = interv.any.year.lower, upp = interv.any.year.upper)]

pdat  <- rbind(pdat1, pdat2)

ggplot(pdat, aes(x = coverage, y = est, group = var, color = var)) +
  theme_bw() +  
  scale_x_continuous(name = "Annual alcohol screening rate", labels = scales::percent, breaks = c(0,0.029,0.1,0.25,0.5,0.75)) + 
  scale_y_continuous(name = "", labels = scales::percent) +
  geom_line(size = 1, linetype = 2) + geom_point(size = 2) +
  geom_errorbar(aes(ymin = low, ymax = upp), size = 1) +
  scale_color_manual(name = "", values = colors.3.green[c(1,3)]) +
  theme(legend.position = "bottom")

ggsave("Figure 3_screening and intervention.png", width = 7, height = 4)
ggsave("Fig 3.tiff", width = 7, height = 4, dpi = 300, unit = "in")

##  4 and 5) APC by groups
#   .........................................

##  APC
pdat <- copy( data[sex != "total" & age != "15-99" & ses != "all" & scenario %in% c(0,2,4,5),.(coverage.lab = paste0(coverage*100,"%"), year, sex, age, ses, PE = APC, LOW = APC.lower, UPP = APC.upper)] )
pdat[, coverage.lab := factor(coverage.lab, levels = c("2.9%","10%","50%", "75%"), ordered = T)]

ggplot(pdat[sex == "female"], aes(x = year, y = PE, fill = coverage.lab)) + 
  ggtitle("APC among women in Germany given different screening scenarios") +
  facet_wrap(age ~ ses, scales = "free", ncol = 3) +
  theme_minimal(base_size = 7) + 
  geom_ribbon(aes(ymin = LOW, ymax = UPP), alpha = 0.5) +
  geom_line() +
  scale_fill_manual(name = "", values = c8Panel_c[c(8,6,4,2)]) + 
  scale_y_continuous(name = "Alcohol per capita consumption") +
  theme(legend.position = "bottom", axis.text.x = element_text(angle = 45, vjust = 0.5, hjust=0.5))

ggsave("Figure 4_APC females.png", width = 6, height = 6)
ggsave("Fig 4.tiff", width = 6, height = 6, dpi = 300, units = "in")

ggplot(pdat[sex == "male"], aes(x = year, y = PE, fill = coverage.lab)) + 
  ggtitle("APC among men in Germany given different screening scenarios") +
  facet_wrap(age ~ ses, scales = "free", ncol = 3) +
  theme_minimal(base_size = 7) + 
  geom_ribbon(aes(ymin = LOW, ymax = UPP), alpha = 0.5) +
  geom_line() +
  scale_fill_manual(name = "", values = c8Panel_c[c(8,6,4,2)]) + 
  scale_y_continuous(name = "Alcohol per capita consumption") +
  theme(legend.position = "bottom", axis.text.x = element_text(angle = 45, vjust = 0.5, hjust=0.5))

ggsave("Figure 5_APC males.png", width = 5, height = 5, dpi = 300, units = "in")
ggsave("Fig 5.tiff", width = 6, height = 6, dpi = 300, units = "in")

##  6) APC sensitivity analyses
#   .........................................

add1 <- copy(data[sex == "total" & age == "15-99" & ses == "all"])
add2 <- copy(sens.data[sex == "total" & age == "15-99" & ses == "all"])
add1$source <- "Main analyses"
add2$source <- "Sensitivity analyses"

pdat <- rbind(add1, add2)
pdat <- pdat[,.(coverage.lab = paste0(coverage*100,"%"), year, sex, age, ses, source, PE = APC, LOW = APC.lower, UPP = APC.upper)]
pdat[, coverage.lab := factor(coverage.lab, levels = c("0%","2.9%","10%","25%","50%", "75%"), ordered = T)]
pdat$coverage.lab 

ggplot(pdat, aes(x = year, y = PE, fill = source)) + 
  ggtitle("APC in Germany in main and sensitivity analyses\ngiven different screening scenarios") +
  facet_grid(coverage.lab ~ ., scales = "free") +
  theme_minimal() + 
  geom_ribbon(aes(ymin = LOW, ymax = UPP), alpha = 0.5) +
  geom_line() +
  scale_fill_manual(name = "", values = c8Panel_c[c(8,2)]) + 
  scale_y_continuous(name = "Alcohol per capita consumption") +
  theme(legend.position = "bottom")

ggsave("Figure 6_sens analyses.png", width = 6, height = 7)
ggsave("Fig 6.tiff", width = 5, height = 6, dpi = 300, units = "in")


#	==========================================================================================================================================
#	==========================================================================================================================================
#	==========================================================================================================================================
#	==========================================================================================================================================
#	==========================================================================================================================================
#	==========================================================================================================================================


#	___________________________________________
#	TABLES
#	___________________________________________

##  APPENDIX TABLE 1 - Descriptives
#   .........................................

tab1 <- copy(data[scenario == 0 & year == 2009 & sex != "total" & age != "15-99" & ses != "all",.(sex, age, ses, 
                                                 LA, LA.lower, LA.upper,
                                                 CD, CD.lower, CD.upper,
                                                 HED, HED.lower, HED.upper,
                                                 DL, DL.lower, DL.upper,
                                                 RISK.MID, RISK.MID.lower, RISK.MID.upper,
                                                 RISK.HIGH, RISK.HIGH.lower, RISK.HIGH.upper)])[order(sex,age,ses)]
tab1 <- merge(tab1, phc.data, by = c("sex","age","ses"))

var.select <- c("LA","LA.lower","LA.upper","CD","CD.lower","CD.upper","HED","HED.lower","HED.upper",
                "RISK.MID","RISK.MID.lower","RISK.MID.upper","RISK.HIGH","RISK.HIGH.lower","RISK.HIGH.upper")

tab1[, (var.select) := lapply(.SD, function(x){x * 100}), .SDcols = var.select]

var.select <- c(var.select,"DL","DL.lower","DL.upper")

tab1[, (var.select) := lapply(.SD, format, digits = 0, nsmall = 1), .SDcols = var.select]

tab1[, LA.out := paste0(LA, "% (",LA.lower," to ", LA.upper,"%)")]
tab1[, CD.out := paste0(CD, "% (",CD.lower," to ", CD.upper,"%)")]
tab1[, HED.out := paste0(HED, "% (",HED.lower," to ", HED.upper,"%)")]
tab1[, DL.out := paste0(DL, " (",DL.lower," to ", DL.upper,")")]
tab1[, RISK.MID.out := paste0(RISK.MID, "% (",RISK.MID.lower," to ", RISK.MID.upper,"%)")]
tab1[, RISK.HIGH.out := paste0(RISK.HIGH, "% (",RISK.HIGH.lower," to ", RISK.HIGH.upper,"%)")]

write.csv(tab1[,.SD,.SDcols = names(tab1)[names(tab1) %like% "sex|age|ses|.out$"]], "S1 table_descriptives.csv", row.names = F)


##  TABLE 2 & 3 - MAIN OUTPUT TABLES
#   .........................................

diff.scen1 <- FUN_DIFF_(aComparator = 1)
diff.scen2 <- FUN_DIFF_(aComparator = 2)
diff.scen3 <- FUN_DIFF_(aComparator = 3)
diff.scen4 <- FUN_DIFF_(aComparator = 4)
diff.scen5 <- FUN_DIFF_(aComparator = 5)

diff.dat    <- rbind(diff.scen1, diff.scen2, diff.scen3, diff.scen4, diff.scen5)
tab  <- dcast(diff.dat, sex + age + ses ~ comparator,value.var = c("HED","DL"))
tab[, order := ifelse(sex == "total" & age == "15-99" & ses == "all", 1,
                       ifelse(sex == "female" & age == "15-99" & ses == "all", 2,
                              ifelse(sex == "male" & age == "15-99" & ses == "all", 3,
                                     ifelse(sex == "total" & age == "15-34" & ses == "all", 4,
                                            ifelse(sex == "total" & age == "35-49" & ses == "all", 5,
                                                   ifelse(sex == "total" & age == "50-64" & ses == "all", 6,
                                                          ifelse(sex == "total" & age == "65-99" & ses == "all", 7,
                                                                 ifelse(sex == "total" & age == "15-99" & ses == "low", 8,
                                                                        ifelse(sex == "total" & age == "15-99" & ses == "mid", 8, 9)))))))))]  

tab2<- tab[order(order),.(sex,age,ses,DL_1,DL_2,DL_3,DL_4,DL_5)]
write.csv(tab2, "table2_reduction in drinking levels.csv", row.names = F)
tab3<- tab[order(order),.(sex,age,ses,HED_1,HED_2,HED_3,HED_4,HED_5)]
write.csv(tab3, "table3_reduction in HED.csv", row.names = F)


#	==========================================================================================================================================
#	==========================================================================================================================================
#	==========================================================================================================================================
#	==========================================================================================================================================
#	==========================================================================================================================================
#	==========================================================================================================================================
```
